# Development and Performance Assessment of a Novel Plasma p-Tau181 Assay Reflecting Tau Tangle Pathology in Alzheimer’s Disease

**DOI:** 10.1101/2023.09.15.23295595

**Authors:** Kenji Tagai, Harutsugu Tatebe, Sayo Matsuura, Zhang Hong, Naomi Kokubo, Kiwamu Matsuoka, Hironobu Endo, Asaka Oyama, Kosei Hirata, Hitoshi Shinotoh, Yuko Kataoka, Hideki Matsumoto, Masaki Oya, Shin Kurose, Keisuke Takahata, Masanori Ichihashi, Manabu Kubota, Chie Seki, Hitoshi Shimada, Yuhei Takado, Kazunori Kawamura, Ming-Rong Zhang, Yoshiyuki Soeda, Akihiko Takashima, Makoto Higuchi, Takahiko Tokuda

## Abstract

Several blood-based assays for phosphorylated tau (p-tau) have been developed to detect brain tau pathologies in Alzheimer’s disease (AD). However, plasma p-tau measured by currently available assays is influenced by brain amyloid and, therefore, could not accurately reflect brain tau deposits. Here, we devised a novel immunoassay that can quantify N- and C-terminally truncated p-tau fragments (mid-p-tau181) in human plasma. We measured plasma p-tau181 levels in 164 participants who underwent both amyloid and tau positron emission tomography (PET) scans using mid-p-tau181 and conventional p-tau181 assays. The mid-p-tau181 assay displayed stronger correlations with tau PET accumulation than the conventional assay in the AD continuum and accurately distinguished between tau PET-positive and -negative cases. Furthermore, the mid-p-tau181 assay demonstrated a trajectory similar to tau PET alongside cognitive decline. Consequently, our mid-p-tau181 assay could be useful in evaluating the extent of brain tau burden in AD.

## Introduction

Estimates project that the number of people with dementia worldwide will reach 153 million by 2050^1^, highlighting the urgent need to address the remarkable health and social impacts of this condition. Alzheimer’s disease (AD), the most prevalent form of dementia, has been characterized as the deposit of two abnormal proteins, namely amyloid-β (Aβ) and tau, in the brain. Over the years, numerous disease-modifying therapies (DMTs) targeting these abnormal proteins have been developed^2^, with the Food and Drug Administration (FDA) having recently approved several DMTs targeting Aβ due to their efficacy in decreasing brain amyloid deposition and modestly slowing AD progression^3–5^. Researchers anticipate that these DMTs will be increasingly utilized in clinical practice, with the potential for developing additional DMTs targeting not only Aβ but also tau in the near future^6^.

Biomarkers have become an integral part of clinical trials on DMTs given their widespread use for subject recruitment and outcome measurements^2^. The amyloid/tau/neurodegeneration (A/T/N) biomarker classification system had been proposed for the biological staging of AD^7^. In particular, positron emission tomography (PET) has emerged as a well-established imaging biomarker for assessing the accumulation of Aβ and tau in the brain^8^, playing a critical role in evaluating novel DMTs^4,9^. The noteworthy decline in amyloid accumulation based on PET findings^10^ may have expedited the FDA approval of aducanumab^2^. Moreover, incorporating tau PET as a screening measure, assuming that high tau accumulation may cause resistance to anti-amyloid therapy, may have played a part in the success of the donanemab clinical trial^4,11^. Despite their usefulness, PET scans are not ideal for frequent assessment or large-scale screening given their limited availability, high cost, and radiation exposure^12^. Similarly, Aβ and p-tau in the cerebrospinal fluid (CSF) have also been accepted as standard biomarkers for qualifying AD pathology^13,14^. However, CSF testing has seen somewhat limited use considering its invasiveness, low-throughput nature, and need for expertise during CSF sampling^12,15^. Based on the present circumstances surrounding the development of biomarkers for AD, one of the most critical unmet needs has been the identification of blood-based biomarkers closely correlated with tau PET parameters that reflect brain tau burden, especially in the context of selecting suitable patients for anti-amyloid therapies.

Recent advances in measurement methodologies have made it possible to quantify minute amounts of proteins in the peripheral blood that are associated with brain diseases, thereby enabling the feasibility of blood-based biomarkers^15^. Notably, there is growing evidence for the clinical significance of plasma phosphorylated tau (p-tau) assays in detecting AD pathology ^16–19^, similar to CSF biomarkers. The C-terminal portion of the tau protein, which predominantly constitutes neurofibrillary tangles^20^, is seldom detectable in biofluids^21–23^. Most of the current immunoassays for plasma p-tau, therefore, employ a pair of antibodies, one of which recognizes the phosphorylated residues, such as Thr181, Thr217, and Thr231, in the mid-portion, while the other identifies the N-terminal region of the tau protein. Accordingly, currently available immunoassays for plasma p-tau detect C-terminally truncated p-tau containing the N-terminus to the mid-domain (N-p-tau) ^12^. Plasma p-tau levels quantified using these N-p-tau assays can accurately differentiate between AD pathology and other tauopathies with high diagnostic accuracy^17,18,24,25^ and help predict future cognitive decline from prodromal phases^26,27^. However, one of the significant problems in the clinical use of N-p-tau assays is their inability to be considered as a surrogate marker for tau burden in the brain^28^ given that the measurements obtained had a more pronounced association with amyloid PET than with tau PET^29–32^. Therefore, the need for developing blood-based tau biomarkers strongly correlated with PET-detectable tau accumulations in the brain remains unmet.

The present study aimed to formulate a novel p-tau assay that can be interchangeable with the tau PET study. Considering that the truncation of the N-terminal of the tau protein may occur subsequent to the truncation of its C-terminal with the formation of neurofibrillary tangle^33^, we hypothesized that the levels of both N- and C-terminally truncated p-tau181 fragments could be correlated with the abundance of tau aggregates in the brain. Thus, we developed a novel mid-region-directed p-tau181 assay by modifying the formerly established N-p-tau181 assay^16^. To validate our hypothesis, we collected plasma samples from subjects who underwent PET with both ^11^C-Pittsburgh Compound B (^11^C-PiB) and ^18^F-florzorotau (aka PM-PBB3/APN-1607) for the examination of Aβ and tau depositions in the brain, respectively.^34,35^ We demonstrated a strong correlation of between plasma levels of p-tau181 fragments measured using our mid-region-directed assay (hereinafter called mid-p-tau181) and tau PET tracer retention but no linear correlation between plasma N-p-tau181 levels and tau PET data, suggesting the clinical usefulness of the novel assay as a surrogate biomarker for PET-visible tau pathologies.

## Results

### Validation of the newly developed mid-p-tau181 assay

The plasma mid-p-tau181 assay exhibited high analytical performance (Supplementary Methods and Results, Supplementary Tables 1–3, Supplementary Figures 1–5), with high precision within and between runs. Spike recovery experiments and parallelism of serially diluted plasma samples confirmed the reliability of the measurements.

### Demographic data

Table 1 summarizes the demographic information of the participants. All participants (n = 164) underwent neuropsychological assessment, including the Clinical Dementia Rating (CDR) scale, Mini-Mental State Examination (MMSE), and frontal assessment battery (FAB), simultaneous amyloid and tau PET imaging, and blood sampling on the day of the PET examination. Cognitively normal (CN) individuals exhibiting negative results on amyloid and tau PET imaging were designated into the CN cohort. Patients with mild cognitive impairment (MCI) and AD who had positive amyloid PET findings were categorized into the AD continuum group (dubbed AD group). Furthermore, subjects with progressive supranuclear palsy (PSP) and other frontotemporal lobar degeneration (FTLD) who had negative amyloid PET findings were classified into the PSP and FTLD cohorts, respectively. No significant differences in age, years of schooling, and gender were observed among the groups. All patient groups showed lower MMSE (CN, 29.3 ± 1.0; AD, 21.9 ± 4.1; PSP, 24.8 ± 5.8; FTLD, 23.8 ± 6.1) and FAB scores (CN, 16.7 ± 1.2; AD, 13.0 ± 3.0; PSP, 12.0 ± 3.6; FTLD, 11.0 ± 4.8) than the CN group (*p* < 0.05, Table 1). Notably, 90% of the AD group comprised subjects with early-stage AD who had a CDR score of 0.5 or 1 accounted; however, the mean MMSE scores of this group were lower than those of the PSP group (*p* < 0.05, Table 1) but did not significantly differ from those of the FTLD group.

**Table 1.** Demographic and blood biomarker data of the participants.

|  | CN | AD | PSP | FTLD |
| --- | --- | --- | --- | --- |
| Demographics |  |  |  |  |
| Number | 40 | 48 | 50 | 26 |
| Age | 66.0 (10.4) | 69.3 (11.4) | 71.2 (7.5) | 65.0 (11.4) |
| Gender<br>(male/female) | 21/19 | 27/21 | 21/29 | 18/8 |
| Years of schooling | 14.8 (1.6) | 13.9 (2.2) | 13.6 (2.6) | 14.0 (2.8) |
| MMSE | 29.3 (1.0)<sup>†</sup> | 21.9 (4.1)<sup>*</sup> | 24.8 (5.8)<sup>*†</sup> | 23.8 (6.1)<sup>*</sup> |
| FAB | 16.7 (1.2)<sup>†</sup> | 13.0 (3.0)<sup>*</sup> | 12.0 (3.6)<sup>*</sup> | 11.0 (4.8)<sup>*</sup> |
| CDR (0.5/1/2/3) | N/A | 23/20/4/1 | N/A | N/A |
| PSPRS | N/A | N/A | 38.1 (18.2) | N/A |
| Blood biomarkers |  |  |  |  |
| Aβ42/40 | 0.093 (0.020)<sup>†</sup> | 0.067 (0.020)<sup>†</sup> | 0.096 (0.048)<sup>†</sup> | 0.089 (0.022)<sup>†</sup> |
| N-p-tau181<br>(pg/mL) | 1.82 (0.79)<sup>†</sup> | 4.07 (1.52)<sup>*</sup> | 2.27 (1.06)<sup>†</sup> | 2.25 (1.25)<sup>†</sup> |
| mid-p-tau181<br>(pg/mL) | 0.83 (0.65)<sup>†</sup> | 2.30 (1.31)<sup>*</sup> | 1.56 (0.91)<sup>*</sup> | 0.96 (0.63)<sup>†</sup> |
| NfL<br>(pg/mL) | 19.5 (12.5)<sup>†‡</sup> | 33.2 (21.3)<sup>*‡</sup> | 58.3 (39.7)<sup>*†</sup> | 51.8 (39.3)<sup>*</sup> |

CN, cognitively normal; AD, Alzheimer's disease; PSP, progressive supranuclear palsy; FTLD, frontotemporal degeneration; MMSE, Mini-Mental State Examination; FAB, Frontal Assessment Battery; CDR, Clinical Dementia Rating scale; PSPRS, progressive supranuclear palsy rating scale; N/A, not applicable; Aβ, amyloid beta; N-p-tau181, phosphorylated tau181 measured using the commercial kit directed to the C-terminally truncated N-terminal fragment of p-tau (Simoa pTau-181 V2.1 Assay, Quanterix); mid-p-tau181, phosphorylated tau181 measured using the originally developed immunoassay directed to both the N- and C-terminally truncated p-tau181 fragments; NfL, neurofilament light chain.
Values are presented as mean ± standard deviation.
<sup>\*</sup>, Significant difference between CN, <sup>†</sup>, between AD, <sup>‡</sup>, between PSP, $P < 0.05$ (corrected by Dunn's multiple comparisons)

### Significant linear correlation between plasma mid-p-tau181 but not N-p-tau181 with tau PET findings in patients with AD continuum

We found a significant linear correlation between the plasma mid-p-tau181 levels and tau PET tracer retention in patients with AD continuum (Figure 1). Voxel-wise analyses revealed a positive correlation between plasma mid-p-tau181 levels and tracer binding in the temporal and parietal cortices [Figure 1A; p < 0.05, corrected for family-wise error (FWE)]. This finding was corroborated by the following three different quantitative indices based on a region of interest (ROI): (1) standardized uptake value ratio (SUVR)^36^ in the temporal meta-ROI that characterizes tau lesions in AD^37^; (2) AD tau score, which is a machine learning-based measure indicating AD-related features of tau PET images^35^; and (3) Braak staging SUVRs based on the neuropathological hypothsesis^36^ (Supplementary Figure 6). These AD signature ROI analyses revealed significant linear correlations between plasma mid-p-tau181 levels and temporal meta-ROI SUVRs (r = 0.506; p = 0.0003), AD tau scores (r = 0.556; p = 0.0003), and SUVRs in the Braak stage III/IV limbic (r = 0.403; p = 0.003) and V/VI neocortical (r = 0.508; p = 0.0003) but not Braak stage I/II entorhinal (r = 0.216; p = 0.149) ROIs. These findings clearly demonstrate that plasma mid-p-tau181 levels could accurately reflect AD-related tau accumulation that spreads from the entorhinal cortex first to the inferior temporal lobe and then to the parieto-occipital regions of the neocortex but not tau accumulation in the Braak I/II region.

**Figure 1.**
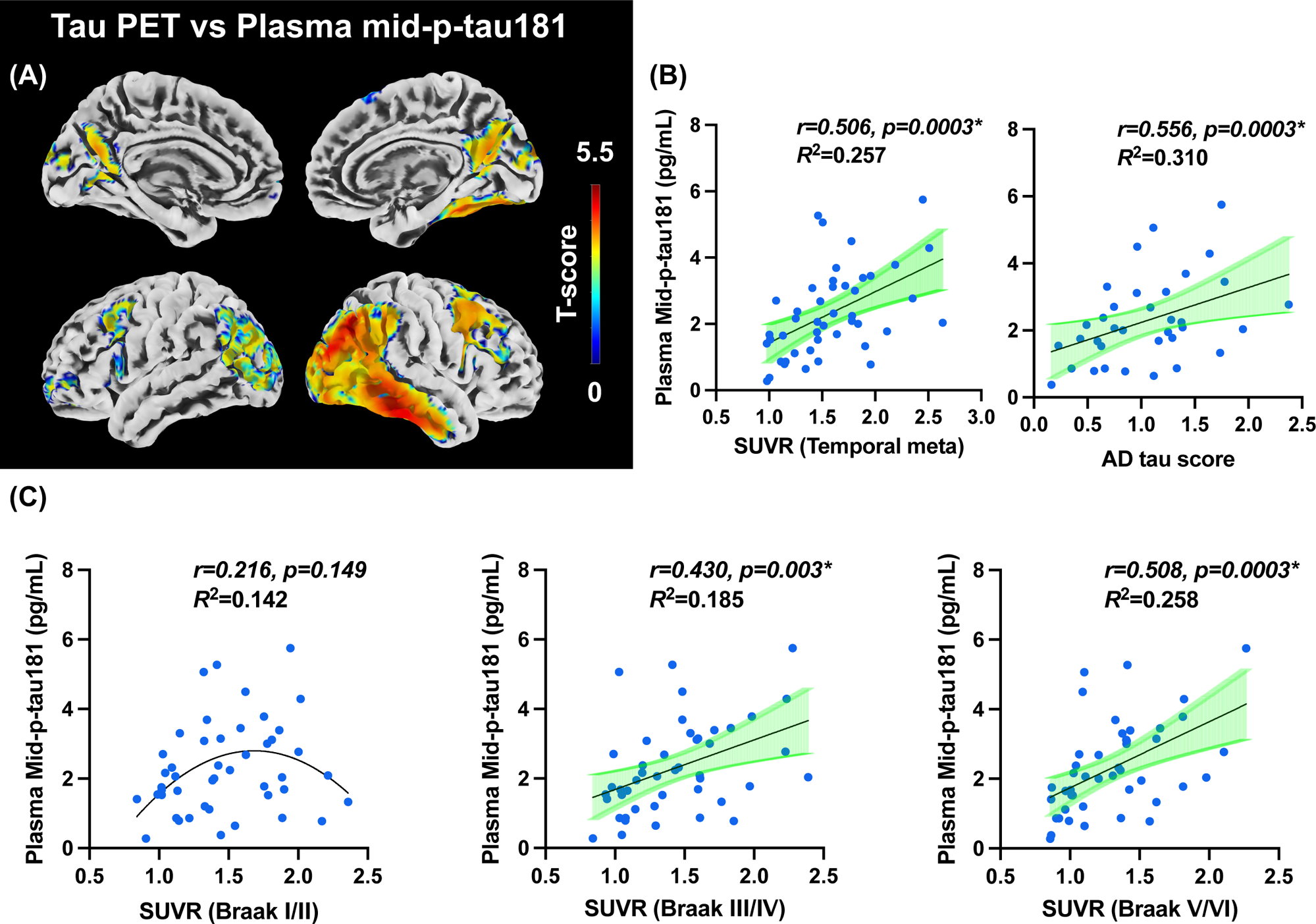
Correlations between plasma mid-p-tau levels and tau PET results in the subjects with AD continuum. (A) The correlation between plasma mid-p-tau181 levels and ^18^F-florzorotau tau PET is depicted through its topographical representation (p < 0.05, family-wise error corrected). (B, C) The correlation between plasma mid-p-tau181 levels and tau PET tracer accumulation in each ROI is portrayed via scatter plots. Pearson’s correlation analysis was employed to calculate the r and p values. Statistical significance was established at p < 0.0125, corrected for multiple comparisons using the Bonferroni method based on the number of ROIs. Regression analysis, indicated by a straight or curved line, depicts the preferred model, with its goodness of fit quantified using the R^2^ value. SUVR, standardized uptake value ratio; AD tau score, a machine learning-based measure indicating AD-related features in tau PET images. Mid-p-tau181, phosphorylated tau181 measured using the originally developed immunoassay directed to both N- and C-terminally truncated p-tau181 fragments.

Conversely, the conventional N-p-tau181 assay (Simoa pTau-181 Advantage V2.1 kit, Quanterix, MA, USA) showed no significant linear correlations with tau PET tracer retention (Figure 2). Voxel-based analysis revealed a weak correlation between these plasma and imaging parameters in the temporal cortex (Figure 2A; p < 0.05, uncorrected); however, no significant correlation was observed upon correction for multiple comparisons (p < 0.05, FWE-corrected). Moreover, none of the three ROI-based analyses revealed significant linear correlations between N-p-tau181 levels and PET parameters (p > 0.05 after Bonferroni’s correction; Figure 2B, C). Instead, these examinations showed inverse U-shaped nonlinear correlations between plasma N-p-tau181 levels and tau PET SUVRs in the temporal meta-ROI (*R*^2^ = 0.402), AD tau score (*R*^2^ = 0.147), and Braak stage III/IV (*R*^2^ = 0.307) and V/VI ROIs (*R*^2^ = 0.223) but not Braak stage I/II ROIs (Figure 2B, C).

**Figure 2.**
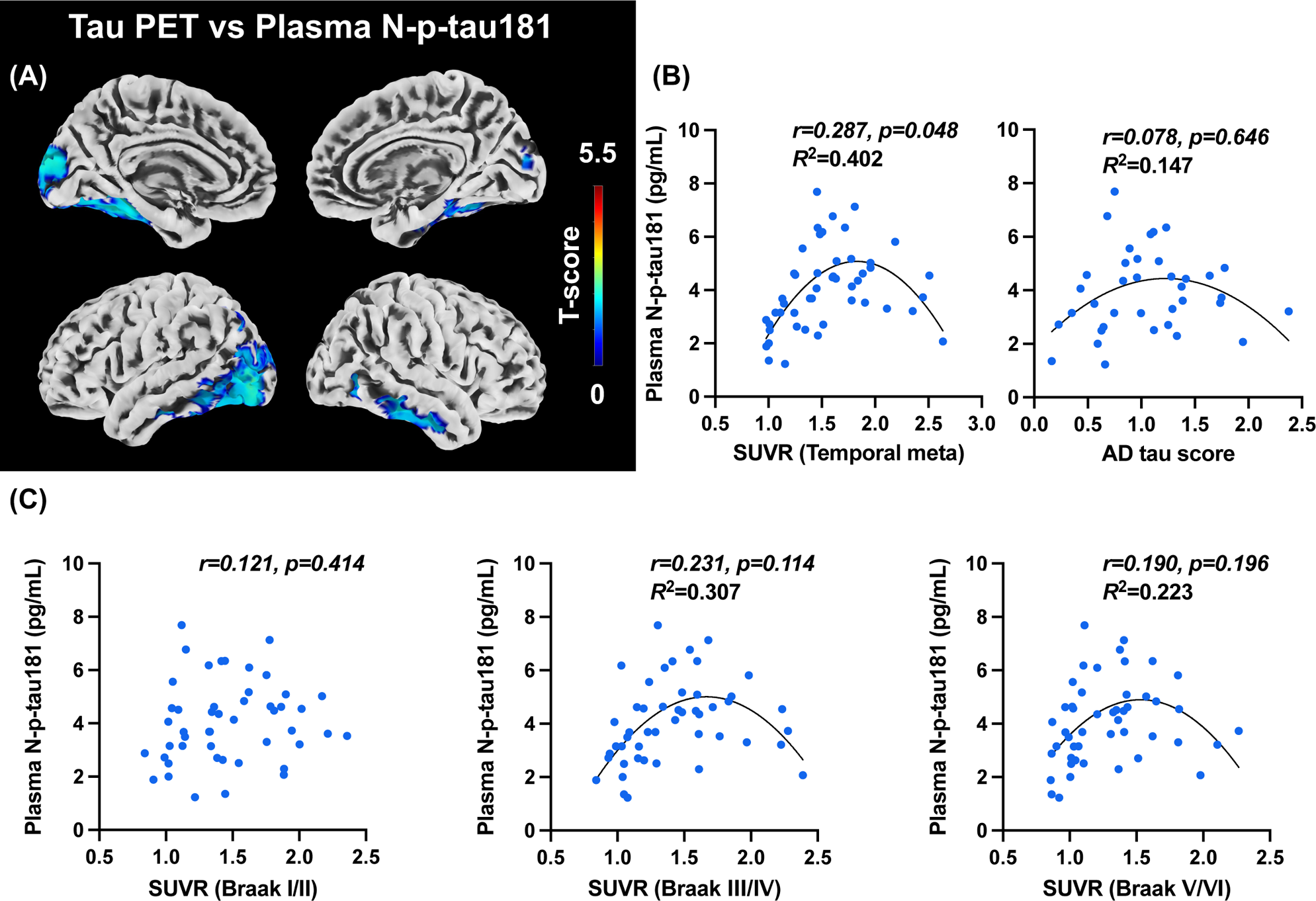
Correlation between plasma N-p-tau levels measured using a conventional p-tau assay and tau PET results in the subjects with AD continuum. (A) The correlation between plasma N-p-tau181 levels and tau PET results is demonstrated through its topographical representation (p < 0.05, uncorrected). (B, C) The correlation between plasma N-p-tau181 levels and tau PET tracer accumulation in each ROI is depicted via scatter plots. Pearson’s correlation analysis was utilized to calculate the r and p values. The preferred model, indicated by regression analysis, is depicted by a straight or curved line, with its goodness of fit quantified using the R^2^ value. SUVR, standardized uptake value ratio; AD tau score, a machine learning-based measure indicating AD-related features in tau PET images. N-p-tau181, phosphorylated tau181 measured using the Simoa pTau-181 V2.1 Assay kit (Quanterix).

### Specific association of plasma mid-p-tau181 with tau PET measures of pathology over amyloid PET measures

Next, plasma p-tau181 levels determined by either the pre-established (N-p-tau181) or newly developed (mid-p-tau181) assay was assessed for association with amyloid-PET and tau-PET measures of pathology (Figure 3). The analysis included both AD and CN groups to avoid the ceiling effect of amyloid PET accumulation that would be the case when analyzing only the subjects with AD continuum. The results of the correlation analysis showed that plasma mid-p-tau181 levels were more strongly associated with tau PET accumulation than with amyloid, while plasma N-p-tau181 levels were more strongly associated with amyloid PET accumulation than with tau (Figure 3A). Multiple linear regression analysis also revealed that plasma mid-p-tau181 levels were significantly influenced only by tau PET accumulation (Amyloid PET: p = 0.056; Tau PET: p < 0.0001, Adjusted R^2^ = 0.450). Meanwhile, plasma N-p-tau181 levels were influenced by both amyloid and tau PET accumulation (Amyloid PET: p = 0.002; Tau PET: p = 0.020, Adjusted R^2^ = 0.420). (See Figure 3B and Supplementary Table 4)

**Figure 3.**
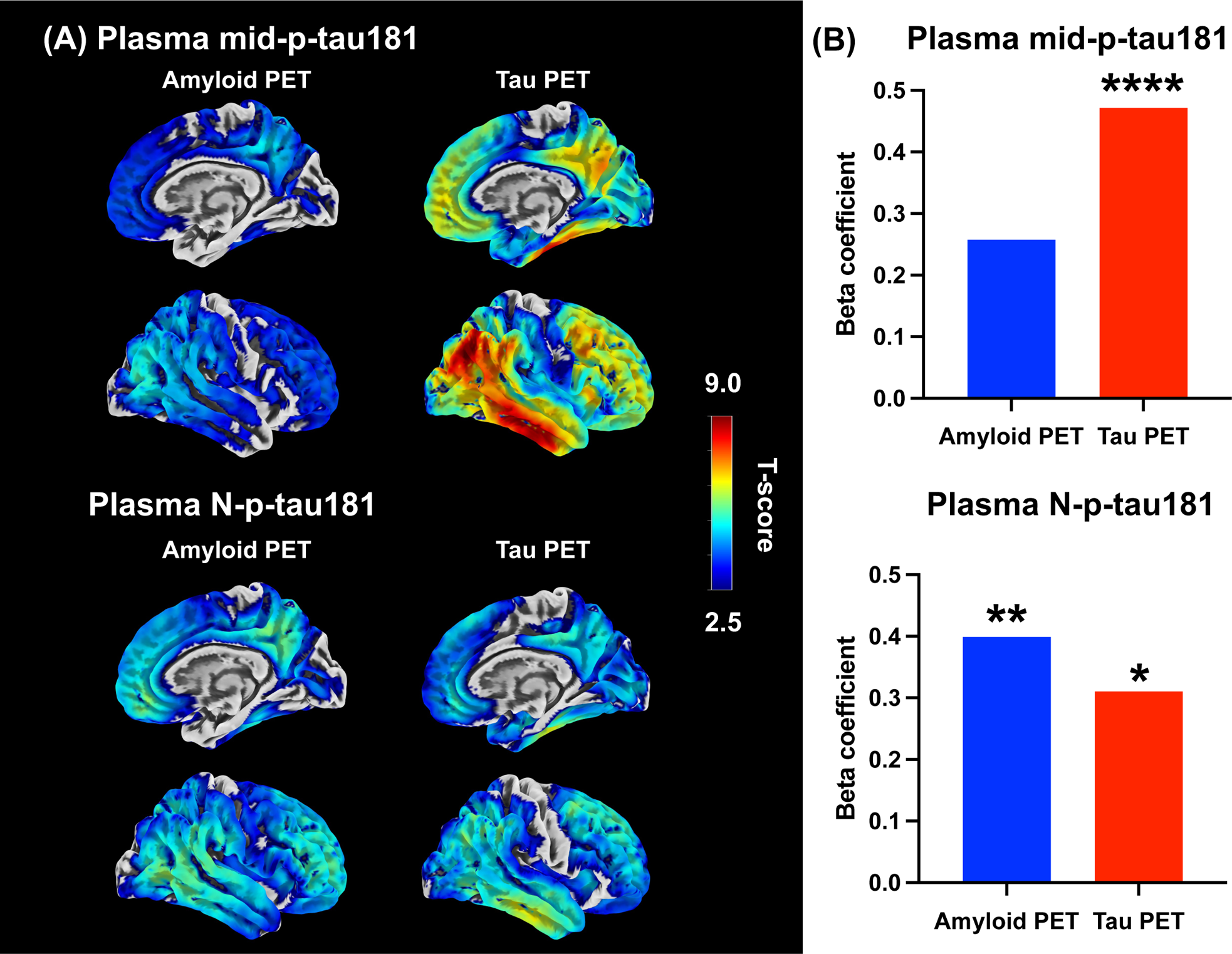
Comparative assessment of Mid-p-tau and N-p-tau levels with amyloid and tau PET within the cognitively normal and AD continuum subjects. (A) The correlation between each plasma p-tau181 level and amyloid and tau PET results in a cohort with CN and AD continuum is demonstrated through its topographical representation (p < 0.05, family-wise error corrected). Upper two rows: associations between mid-p-tau and amyloid-PET (left column) and tau-PET (right column); Lower two rows: associations between N-p-tau and amyloid-PET (left column) and tau-PET(right column). (B) The bar graphs illustrate beta coefficient values of amyloid and tau PET in multiple liner regression analysis with each p-tau assay. Statistically significant differences are denoted as follows: p < 0.0005 (****), p < 0.005 (**), p < 0.05 (*). P-values were corrected for multiple comparisons using Bonferonni correction with the number of explanatory variables.

### Comparisons between the two plasma p-tau181 assays according to their correlation with structural imaging biomarkers

The relationship of brain volume and cortical thickness, assessed by magnetic resonance imaging (MRI), with measures of each p-tau181 assay was also examined. Voxel-based morphometry revealed significant negative correlations between the plasma mid-p-tau181 concentration and gray matter volume in the neocortex, primarily in the temporal and parietal areas (Figure 4). The ROI-based analysis also confirmed a correlation between mid-p-tau181 and cortical thinning in AD-signature regions defined as described elsewhere^38^ (r = −0.406; p = 0.005). In contrast, N-p-tau181 did not exhibit any correlation with structural imaging measures.

**Figure 4.**
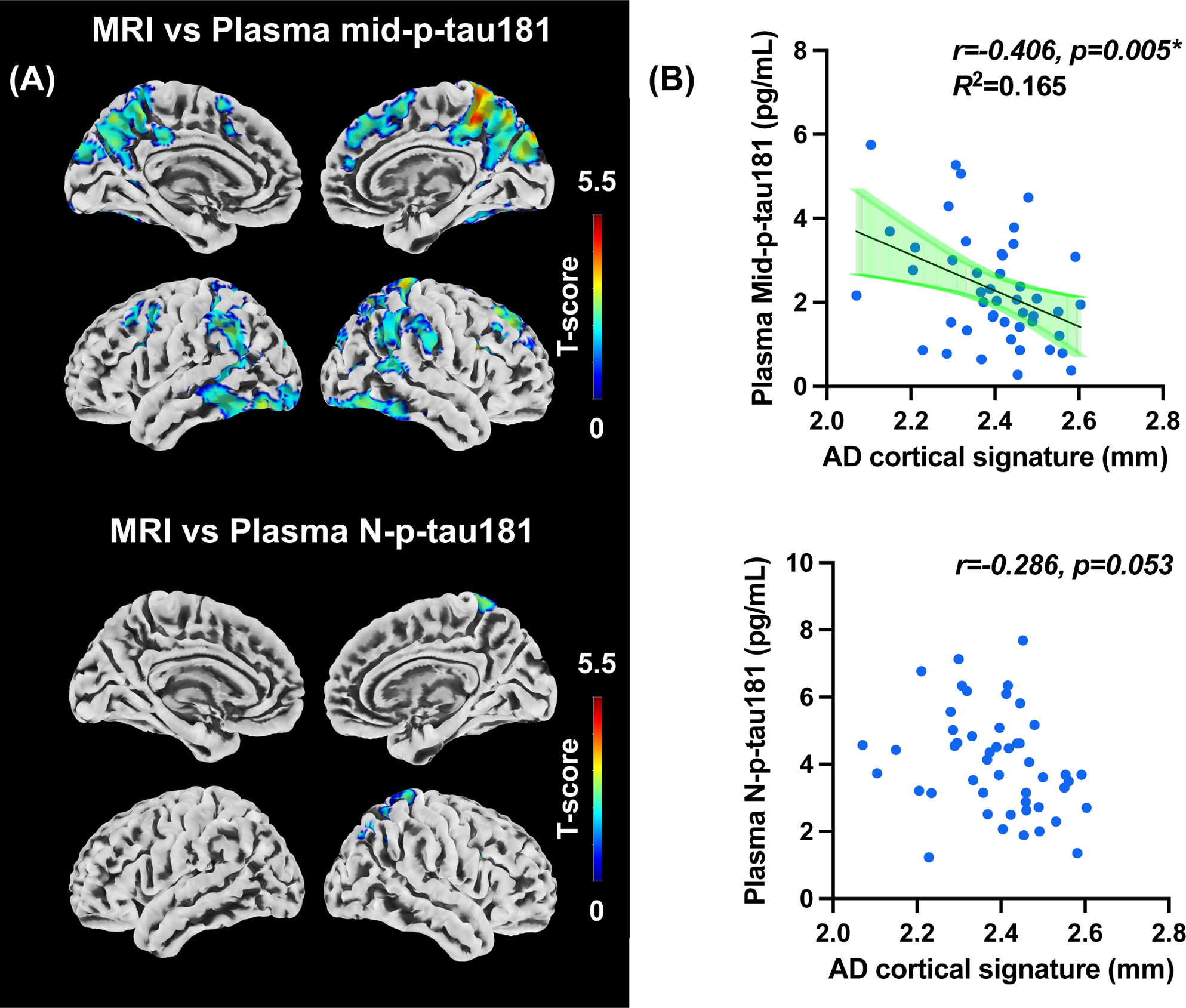
Correlations between plasma mid-p-tau and N-p-tau levels and MRI structural images in the subjects with AD continuum. (A) The correlation between p-tau181 measured using the mid-p-tau181 or N-p-tau assay and brain atrophy is depicted through its topographical representation (p < 0.05, uncorrected). (B) The correlation between plasma mid-p-tau181 and N-p-tau levels measured using each assay and the thinning of the AD cortical signature is demonstrated via scatter plots. Pearson’s correlation analysis was utilized to calculate. N-p-tau181, phosphorylated tau181 measured using the Simoa pTau-181 V2.1 Assay kit (Quanterix); mid-p-tau181, phosphorylated tau181 measured using the originally developed immunoassay directed to both N- and C-terminally truncated p-tau181 fragments.

### Discriminating between tau PET statuses using plasma mid-p-tau181 levels

Using three different ROI-based methods, we qualitatively assessed the PET-detectable tau burden in the CN subjects and AD continuum patients. Thereafter, receiver operating characteristic (ROC) curve analyses were conducted to determine plasma mid-p-tau181 cutoff levels for discriminating between individuals with positive and negative AD-type tau PET findings. The PET finding classification was based on the cutoff values of imaging-based Braak staging, temporal meta-ROI SUVR, and AD tau score. The Braak stage in each subject was determined according to the ROI with the highest Z-score (see Methods for detailed procedures), with stages 0 and I/II indicating tau-negative and stages III/IV and V/VI being classified as tau-positive (Supplementary Table 5). The cutoff values for the meta-ROI SUVR and AD tau score were determined using ROC curve analyses in the current and previous studies^35^, respectively (Supplementary Table 5). Consequently, our findings showed that tau-positive AD cases had significantly higher plasma mid-p-tau181 concentrations than did tau-negative AD and CN cases (Figure 5A; p < 0.0001). Furthermore, area under the ROC curve (AUC) values for the mid-p-tau181-based differentiation between tau PET positivity and negativity defined by all three methods exceeded 0.85 (Table 2, Figure 5B), indicating that this plasma biomarker with reference to PET-based classifications had robust discriminative power.

**Figure 5.**
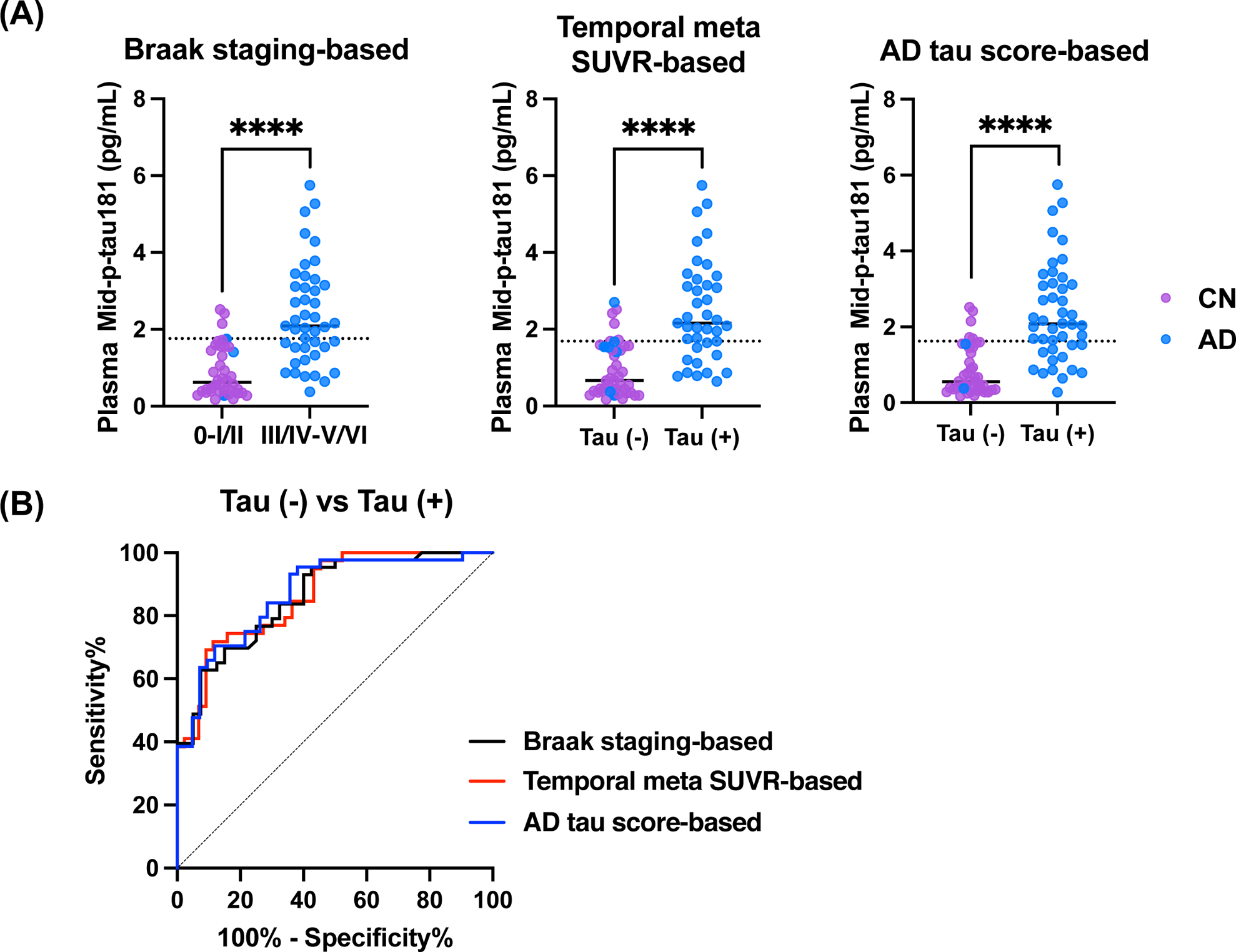
Scatter plots and ROC curves of the mid-p-tau181 showing its ability to discriminate between tau PET statuses determined by three different approaches in the cognitively normal and AD continuum subjects. Scatterplots (A) and ROC curves (B) illustrating the relationship between mid-p-tau181 levels and positive/negative tau PET results as determined by three different methods. In the scatterplot, CN subjects are represented in purple, whereas AD continuum patients are depicted in blue. The dotted line represents the mid-p-tau181 cutoff calculated based on each approach. p < 0.0001 (****), as assessed by Mann–Whitney U test.

**Table 2.** Performance of mid-p-tau181 in discriminating tau PET status determined using three different methods of evaluating brain tau burden on tau PET in the cognitively normal and AD continuum subjects.

|  | Sensitivity (%) | Specificity (%) | Cutoff value (pg/mL) | AUC |
| --- | --- | --- | --- | --- |
| Braak staging | 62.8 | 92.5 | 1.76 | 0.859 |
| Temporal meta SUVR | 71.8 | 88.6 | 1.69 | 0.855 |
| AD tau score | 70.5 | 88.1 | 1.62 | 0.870 |

All parameters were estimated from receiver operating characteristic curve analyses. We set each cutoff value according to Youden index obtained in the respective receiver operating characteristic analyses.
SUVR, standardized uptake value ratio; AUC, area under the receiver operating characteristic curve.

### Discriminating among diagnostic groups and between amyloid PET statuses using plasma biomarkers

Supplementary Figure 7A illustrates group comparisons of four AD-related plasma biomarkers, including the mid-p-tau181 measured using our in-house assay, and the corresponding values for each diagnostic group being presented in Table 1. Our results indicated significant differences in Aβ42/40 and N-p-tau181 concentrations between the AD and other groups. Additionally, the AD group exhibited significantly higher mid-p-tau181 concentrations than did all other groups except the PSP group. Moreover, all disease groups had increased neurofilament light chain (NfL) concentrations compared to the CN group. ROC curve analyses (Supplementary Figure 7B) revealed the following order of the biomarker performances for discriminating between individuals with and without ^11^C-PiB-PET-positive amyloid pathology when all participants were incorporated: N-p-tau181 (AUC = 0.867), Aβ42/40 (AUC = 0.850), mid-p-tau181 (AUC = 0.771), and NfL (AUC = 0.555).

### Trajectory analyses of imaging- and blood-based A/T/N biomarkers along with cognitive decline in the CN and AD continuum subjects

The imaging biomarkers examined in this cohort revealed that the A marker (i.e., amyloid PET index) displayed a sigmoidal trajectory when plotted against cognitive deficits and reached a plateau when the MMSE score was around 20 points, the point at which cognitive decline became apparent (Figure 6A). Conversely, the T marker (i.e., tau PET index) displayed a linearly progressive increase with a decrease in the MMSE scores. The N marker (i.e., the volumetric MRI index) also tended to show a linear increment similar to that for the T marker, albeit with a lesser z-score range.

**Figure 6.**
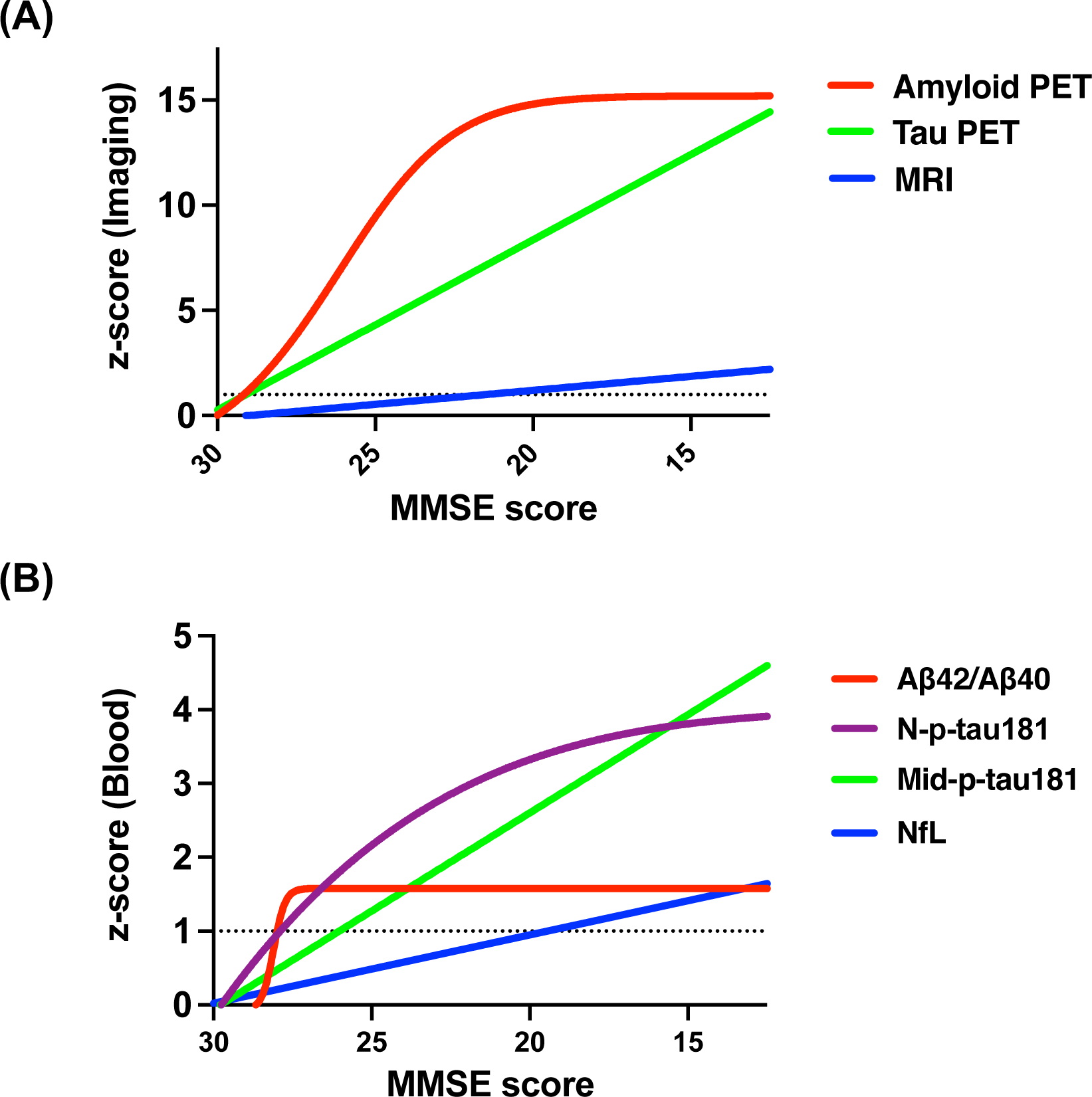
Trajectories of the imaging and plasma A/T/N biomarkers along with the decline in the MMSE scores. Trajectories of the changes in imaging (A) and blood-based (B) A/T/N biomarkers with the decline in MMSE scores. The relationship between MMSE scores of the CN and AD continuum subjects and the z-scores of each biomarker is presented as a regression line that is either straight or sigmoidal, with the best fitting model being selected. The biomarkers were distinguished using red, green, and blue for both imaging and blood-based biomarkers, whereas N-p-tau181 was presented separately in purple. The dotted line indicates z-score = 1.

The blood-based biomarkers displayed trajectories closely resembling those of the corresponding imaging biomarkers, although their dynamic ranges were inferior to those of the imaging biomarkers (Figure 6B). Notably, the two different blood-based T markers showed clearly distinct trajectories such that plasma mid-p-tau181 levels exhibited a linearly progressive increase similar to the tau PET index with the decline in MMSE scores, whereas plasma N-p-tau181 levels displayed a sigmoidal trajectory similar to that for amyloid PET with the decrease in MMSE scores. The A (Aβ42/40) and N (NfL) markers showed a sigmoidal curve and a linear progression, respectively, similar to those for the corresponding imaging biomarkers.

## Discussion

The current study aimed to develop a novel plasma p-tau biomarker, mid-p-tau181, which could accurately reflect the AD-type tau burden in the brain assessed by tau PET, and could thus be interchangeable with tau PET imaging. Notably, our results showed that the plasma mid-p-tau181 exhibited a stronger association with tau PET than amyloid PET in AD brains and was accordingly able to discriminate individuals presenting positivity and negativity for PET-detectable AD tau pathologies. Amyloid depositions in the brain have been known to affect the plasma levels of N-terminally untruncated forms of p-tau181 (N-p-tau181) detected by most conventional p-tau assays.^17,26^ By contrast, plasma mid-p-tau181 levels could have sufficient accuracy in determining the existence of AD-type tau pathology without being affected by amyloid accumulation. Moreover, the plasma mid-p-tau181 levels displayed a linear elevation along with declines in MMSE scores, similar to tau PET measures. These results strongly suggest the potential utility of plasma mid-p-tau181 as a blood-based biomarker for detecting and staging tau pathologies from preclinical to advanced clinical phases of the AD continuum.

Various non-clinical and clinical studies have suggested that an increase in soluble p-tau in biofluids could be strongly associated with Aβ aggregation.^17,29,30,39–42^ This evidence indicates that tau hyperphosphorylation occurs in neurons exposed to Aβ, with p-tau levels increasing in response to Aβ deposition. Consistent with this notion, plasma p-tau is highly valuable for distinguishing AD from other Aβ-negative neurodegenerative diseases.^18,43–45^ In addition, concentrations of N-terminally intact p-tau measured using conventional assays have been recognized as an early marker for AD, given the increase in their values alongside Aβ deposition^17,44,46–49^ rather than PET-visible tau aggregations.^19,28,30^ Meanwhile, tangle maturation also needs to be considered when interpreting the levels of soluble p-tau in biofluids measured using immunoassay methodologies. Most current approaches for quantifying soluble p-tau via immunoassay predominantly target tau forms phosphorylated at positions 181, 217, or 231 and bearing N-terminal epitopes.^12^ The C-terminal portion of the tau protein involved in its aggregation^50^ is rarely present in soluble tau species. Thus, soluble tau species present in the body fluids usually include only the N-terminal-to-mid region epitopes, ensuring consistent quantification using N-p-tau assays currently accessible.^12^ In the meantime, N-terminal and C-terminal cleavages occur in tau proteins insolubilized and deposited in the brain, which has been postulated to play a role in tangle evolution.^33,51–53^ Therefore, conventional plasma N-p-tau assays are likely to be incapable of assessing N-terminally truncated tau species that can be elevated with the advancement of tangle formations. Concurrent with this perspective, a study investigating the associations between various plasma p-tau species and amyloid/tau PET demonstrated that all plasma p-tau species measured by N-p-tau assays were more tightly associated with amyloid-PET than tau-PET.^30^ These findings support the notion that currently available p-tau assays are not interchangeable with tau PET but can provide surrogates for amyloid depositions.

Conversely, our mid-p-tau181 assay strongly correlated with tau but not amyloid PET indices in AD brains. Since ^18^F-florzolotau, which was employed as a radioligand for tau PET imaging here, exerts high sensitivity and specificity for tau versus amyloid aggregates in the AD spectrum,^34^ the plasma mid-p-tau181 measurement for the first time offers the blood-based proper “T” biomarker in the ATN framework, corresponded to the AD-type tau accumulation in tau PET scans.

Moreover, the characteristics of mid-p-tau181 suggest its usefulness as a biomarker for predicting the therapeutic response to current DMTs targeting Aβ. These therapies are more likely to be effective when introduced before substantial tau aggregation.^2,11,54,55^ Our results showed that our plasma mid-p-tau181 could discriminate between tau-positive and -negative subjects with a specificity greater than 85%, and that plasma mid-p-tau levels showed a significant linear correlation with tau PET tracer retention in patients with AD continuum. Therefore, reasonable estimation of brain tau accumulation by using plasma levels of mid-p-tau181 could identify patients with high plasma mid-p-tau levels, in whom brain tau aggregation has been suggested to have progressed beyond the critical point, and could thereby help select patients with low-to-moderate tau burden who are more likely to benefit from DMTs targeting Aβ, like so in the phase 3 study of donanemab.^11^

Plasma mid-p-tau181 levels were also correlated with brain atrophy and increased with the progression of cognitive impairment evaluated using MMSE scores in the trajectory analysis. Meanwhile, plasma N-p-tau181 levels increased in the early stage even when MMSE scores were within the normal range, and its increase slowed down in the later phase when cognitive impairment advanced. In support of these findings, the elevation of CSF p-tau181 measured with an immunoassay targeting the mid-portion of tau protein occurred later than CSF N-p-tau181,^49^ as observed in our plasma assay.

However, the increase in CSF N-p-tau181 species ceased and decreased after neuronal dysfunction started in the familial AD cohort.^56^ In light of these current and previous findings, plasma mid-p-tau181 serves as a biomarker to monitor the disease progression over a phase characterized by fibrillar tau deposition and consequent neurodegeneration and cognitive declines, whereas N-p-tau levels indicate earlier pathologies primarily induced by Aβ accumulations.

The PSP group also showed a significant elevation in plasma mid-p-tau181, deviating from the results of the N-p-tau assay and leading to a slight decrease in specificity to AD. Phosphorylation in the mid-region has also been observed in non-AD tauopathies, such as PSP,^57^ with some previous studies reporting that soluble mid-p-tau levels in CSF might be elevated in non-AD tauopathies.^58^ However, plasma mid-p-tau181 levels were not correlated with tau PET accumulation in the PSP patients (Supplementary Figure 8). Accordingly, the underlying pathophysiological basis for the observed increase in plasma mid-p-tau181 levels in our PSP group remains elusive.

This study has several limitations worth noting. Firstly, our sample size was modest, and longitudinal data were lacking. As such, clinical cohort studies assessing neuroimaging- and fluid-based biomarkers with a larger scale are required to justify the present findings and are underway. Secondly, it is yet to be examined whether plasma mid-p-tau181 correlates with measures of tau PET with other tracers, including ^18^F-flortaucipir and ^18^F-MK6240, in the AD spectrum. ^18^F-florzolotau and these tracers yield similar cortical and limbic maps of AD-type tau aggregates,^59^ while there might be diversity of the compound reactivity with various tau fibril subtypes. Moreover, distinct significances of tau phosphorylation at different sites may need to be taken into account, as a few of the amino acid residues can be phosphorylated at an incipient stage of AD pathology, followed by other epitopes.^56,60^ In addition, previous reports indicated a moderate correlation between plasma p-tau217 and postmortem or PET-detectable tau pathologies in the AD brain,^18,19,32^ and it is accordingly intriguing to compare the performances of mid-p-tau assays targeting phosphorylation sites other than Thr181.

To conclude, the present multimodal bioanalysis of AD-spectrum and control cohort subjects has demonstrated the capability of the newly developed plasma mid-p-tau181 assay for pursuing the dynamics of tau fragments in association with the tangle formation and for the biological staging of AD. Although tau fragments in the CSF have been measured and shown to correlate with tau pathologies,^61,62^ there are still unmet needs for implementing comprehensively validated blood-based biomarkers that accurately reflect cerebral tau burden with minimal interference by amyloid deposition. Our novel mid-p-tau assay potentially offers a biomarker with high accessibility and functionality for evaluating the evolution of neurodegenerative tau pathogenesis that has been investigated only by PET. Notwithstanding the necessity for additional proofs, this technology will also be applicable to the selection of patients most suitable for anti-Aβ DMTs and the examining efficacies of tau-targeted therapies.

## Methods

### Participants

We recruited a cohort of 186 individuals comprising 43 CN individuals, 30 patients with MCI due to AD, 31 patients with dementia due to AD, 52 patients with PSP, and 30 patients with other FTLD syndromes between January 2018 and September 2022. The other FTLDs comprised 12 cases with corticobasal syndrome (CBS), 10 cases with behavioral-variant frontotemporal dementia (BvFTD), 7 cases with frontotemporal dementia and parkinsonism linked to chromosome 17 MAPT (FTDP-17/MAPT), and 1 case of primary progressive aphasia. CN individuals were those aged older than 40 years who had no history of neurological and psychiatric disorders, had a MMSE score of ≥28, or had a Montreal Cognitive Assessment score of ≥26 and Geriatric Depression Scale score of ≤5.^35^ Patients with MCI and dementia due to AD underwent clinical evaluations. Cognitive impairment severity was defined as MMSE < 24 and CDR ≥ 1 for dementia and MMSE ≥ 26 and CDR = 0.5 for MCI.^35^ Patients with PSP and other FTLD syndromes were diagnosed according to established criteria previously reported.^34,35^

In the present study, the diagnosis of patients with MCI and dementia due to AD required brain amyloid positivity in PET scans, and they were combined and categorized into the AD group based on the concept of the AD continuum. We excluded patients with PSP and other FTLD who had positive amyloid PET results to eliminate the influence of mixed pathology. Those with other FTLDs were heterogeneous and categorized into the FTLD group. Furthermore, CN individuals with a positive result on either or both amyloid and tau PET scans were also excluded as they were considered to have been in the preclinical AD stage. Amyloid positivity was defined based on ^11^C-PiB-PET visual inspection performed by a minimum of three specialists with expertise in the field.^34^ Tau PET negativity in CN individuals was defined according to the AD tau score (< 0.1986) and PSP tau score (< 0.3431) as reported previously.^35^ These scores, which were calculated from ^18^F-florzorotau PET images via our established machine learning algorithm, possess a high degree of sensitivity and specificity in discriminating CN individuals from patients with AD and PSP.^35^ Consequently, after excluding 3 CN individuals and 11 MCI, 2 AD, 2 PSP, and 4 FTLD patients according to the aforementioned criteria, our final cohort consisted of 164 participants comprising 40 CN individuals and 48 AD, 50 PSP, and 26 FTLD patients.

This study was approved by the National Institutes for Quantum Science and Technology Certified Review Board. Written informed consent was obtained from all participants and spouses or close family members when participants were cognitively impaired. This study was registered with the UMIN Clinical Trial Registry (UMIN-CRT; number 000041383).

### Blood sampling

We obtained blood samples through venous puncture on the same day as the PET scan. A total of 8 mL of blood was collected in ethylenediaminetetraacetic acid (EDTA)-containing tubes. After collection, plasma was separated by centrifugation for 10 min at 2000*g*, aliquoted into polypropylene tubes, and then stored at −80°C until analysis.

### Measurements of blood biomarkers

We developed a novel immunoassay able to quantify plasma levels of both N- and C-terminally truncated p-tau181 fragments run on a highly sensitive automated digital ELISA platform (Simoa HD-X Analyzer, Quanterix, Lexington, KY, USA) and measured the levels of p-tau181 including such fragments in human plasma. The details of the procedures for method validation of this original immunoassay are described in the Supplementary Methods and Results. We also quantified plasma levels of the A/T/N biomarkers^7^ (Aβ42, Aβ40, p-tau181, and NfL instead of total tau in the original paper) utilizing the Simoa platform (Quanterix) equipped with validated assay kits. Procedures were performed following the manufacturer’s instructions. This study employed the Aβ42/Aβ40 ratio as a proxy for cerebral amyloid burden. Plasma p-tau181 measured using the commercial kit (Simoa pTau-181 V2.1 Assay, Quanterix) was defined as N-p-tau181, whereas plasma p-tau181 measured using the originally developed immunoassay run on the Simoa system was defined as mid-p-tau181.

All plasma samples were diluted four times with the respective sample diluent before the assays to minimize matrix effects. All plasma samples were run in duplicate with the same lot of standards. The relative concentration estimates of plasma biomarkers were calculated according to their respective standard curves.

### PET and MRI data acquisition

Amyloid and tau deposits in the brains of all participants were assessed using PET with ^11^C-PiB and ^18^F-florzorotau as described in other clinical trials (UMIN-CRT; number 000026385, 000026490, 000029608, 000030248, and 000043458). One PSP patient who had already been confirmed to be Aβ-negative at another facility no longer underwent ^11^C-PiB-PET at our center. The scan protocol was described as follows: parametric ^11^C-PiB-PET images were acquired 50–70 min after injection (injected dose: 528.5 ± 65.5 MBq, molar activity 90.2 ± 26.2 GBq/μmol); ^118^F-florzorotau PET images were obtained 90–110 min after injection (injected dose: 186.6 ± 7.4 MBq, molar activity 244.2 ± 86.7 GBq/μmol). PET was primarily conducted using a Biograph mCT flow system (Siemens Healthcare), with some cases using the Discovery MI (GE Healthcare) (9 ^11^C-PiB scans in CN individuals; 22 ^11^C-PiB scans and 3 ^18^F-florzorotau scans in AD patients; 7 ^11^C-PiB scans in PSP patients; and 4 ^11^C-PiB scans in FTLD patients) and an ECAT EXACT HR+ scanner (CTI PET Systems, Inc.) (three ^11^C-PiB scans in CN individuals, four ^11^C-PiB scans in AD patients, nine ^11^C-PiB scans in PSP patients, and three ^11^C-PiB scans in FTLD patients). Acquired PET images were reconstructed using the filtered back projection method with a Hanning filter. MRI examination was conducted simultaneously with PET using a 3-T scanner (MAGNETOM Verio; Siemens Healthcare). The anatomical images were acquired using a three-dimensional T1-weighted gradient echo sequence that produced a gapless series of thin sagittal sections (TE = 1.95 ms, TR = 2300 ms, TI = 900 ms, flip angle = 9°, acquisition matrix = 512 × 512 × 176, voxel size = 1 × 0.488 × 0.488 mm^3^).

### Imaging analyses

All images were preprocessed using PMOD software (version 4.3, PMOD Technologies Ltd), FreeSurfer 6.0 (http://surfer.nmr.mgh.harvard.edu/), MATLAB (The Mathworks, Natick, MA, USA), and Statistical Parametric Mapping software (SPM12, Wellcome Department of Cognitive Neurology). PET images were co-registered with individual anatomical T1-weighted MR images, and SUVR images were generated using each reference region. The cerebellar cortex was the reference region for the ^11^C-PiB-PET images. For ^18^F-florzorotau PET images, an optimized reference region was set through an in-house MATLAB script that considered the distribution of diverse tau lesions throughout the entire gray matter and extracted optimized reference regions on an individual basis.^63^ Each PET and MR image was also normalized to the Montreal Neurologic Institute space using the Diffeomorphic Anatomical Registration Through Exponentiated Lie Algebra (DARTEL) algorithm and was smoothed with a Gaussian kernel at 8-mm full-width at half maximum in voxel-wise analyses.

We performed an ROI analysis targeting AD pathologies on each imaging modality to quantify the regional amyloid/tau burden and cortical thinning. The amyloid burden was assessed using a Centiloid atlas (frontal, temporal, parietal, precuneus, anterior striatum, and insula) implemented in the PMOD Neuro Tool (PMOD Technologies Ltd). Each Centiloid SUVR was calculated and converted to a Centiloid score (CL score)^64^ using PET data from 12 young CN individuals aged 23–43 years and 25 cases of AD patients scanned at our institution. Tau burden was assessed using ROIs targeting tau pathology associated with AD labeling through FreeSurfer, Braak staging ROIs (I/II, III/IV, V/VI),^36^ and temporal meta-ROI (entorhinal, amygdala, parahippocampal, fusiform, inferior temporal, and middle temporal).^37^ We excluded the hippocampus from the Braak stage I/II ROI because of potential spill-in from the choroid plexus.^34^ Additionally, we also estimated the AD tau score to assess AD-type tau burden in the brain, which was calculated using an Elastic Net model trained on tau PET data as previously reported.^35^ A qualitative analysis based on the values obtained from these ROI analyses was also conducted to evaluate the presence of tau lesions. For Braak staging, the SUVR values were converted to z-values based on another young CN cohort, and the highest stage was assigned based on the average regional Z-score (>2.5). Those with stages 0–I/II were classified as tau-negative, whereas those with stages III/IV–V/VI were classified as tau-positive. The cutoff value of temporal meta-ROI SUVR was set at 1.105 to maximize the differentiation between CN individuals and AD patients during ROC analysis (see Supplementary material for detailed information). and the cutoff value of AD tau score was set at 0.1986 as described elsewhere.^35^ Cortical thickness was measured using the cortical signature of AD through FreeSurfer (medial temporal, inferior temporal, temporal pole, angular, superior frontal, superior parietal, supramarginal, precuneus, and middle frontal).^38^

### Statistical analyses

Statistical analyses were conducted using GraphPad Prism version 9 (GraphPad Software) and R version 4.3.1. Initially, group comparisons were performed using the Kruskal–Wallis test or Mann–Whitney U test for demographic data and measured blood biomarker values and Fisher’s exact test for gender (p < 0.05, corrected by Dunn’s multiple comparisons). Subsequently, correlation analyses were conducted to verify the association between each p-tau181 assay and each imaging biomarker. During voxel-based analyses, a linear regression model was applied using SPM12. The extent threshold was established based on the expected voxels per cluster. For multiple voxel comparisons, family-wise error corrections at the cluster level were applied (p < 0.05, FWE-corrected). During ROI-based analyses, Pearson’s correlation analyses were performed (p < 0.05, corrected by Bonferroni multiple comparisons), and nonlinear regression analysis (quadratic) was conducted when no significant correlation was observed. Results were adopted when the nonlinear analysis based on Akaike’s Information Criterion (AIC) showed a better fit than the linear one. Besides, we also incorporated a multiple linear regression analysis to explore the relationships between amyloid, tau, and each plasma pTau level. In addition, ability of each blood biomarker to discriminate between the presence or absence of AD pathology, as defined by amyloid or tau PET positivity, was also evaluated by calculating AUC values from ROC curve analyses. The Youden index maximizing sensitivity plus specificity minus one determined the optimized cutoff value. Finally, to explore the trajectories from CN to AD for each blood/imaging biomarker, we converted each biomarker value to a z-value based on CN data. Thereafter, we examined their relationship with cognitive dysfunction (MMSE score). A linear or sigmoidal 4PL regression analysis was adapted, and the better-fitting model was selected based on AIC.

## Supporting information

Supplementary Methods and Results

## Data availability

The data supporting this study’s findings are available from the corresponding author on reasonable request. Sharing and reuse of data require the expressed written permission of the authors, as well as clearance from the Institutional Review Boards.

## Acknowledgements

The authors thank all patients and their caregivers for participation in this study, as well as clinical research coordinators, PET and MRI operators, radiochemists, and research ethics advisers at QST for their assistance with the current projects. We thank APRINOIA Therapeutics for kindly sharing a precursor of ^18^F-florzorotau. The authors acknowledge support with the recruitment of patients by Shunichiro Shinagawa at the Department of Psychiatry, Jikei University School of Medicine; Shigeki Hirano at the Department of Neurology, Chiba University; Taku Hatano, Yumiko Motoi, and Shinji Saiki at the Department of Neurology, Juntendo University School of Medicine; Ikuko Aiba at the Department of Neurology, National Hospital Organization Higashinagoya National Hospital; Yasushi Shiio and Tomonari Seki at the Department of Neurology, Tokyo Teishin Hospital; Hisaomi Suzuki at the National Hospital Organization Shimofusa Psychiatric Medical Center.

## Author contributions

**Kenji Tagai, Harutsugu Tatebe**: Conceptualization, Methodology, Formal analysis, Writing - original draft. **Sayo Matsuura, Zhang Hong**: Measurement of samples. **Naomi Kokubo, Kiwamu Matsuoka, Hironobu Endo, Asaka Oyama, Kosei Hirata, Hitoshi Shinotoh, Yuko Kataoka, Hideki Matsumoto, Masaki Oya, Shin Kurose, Keisuke Takahata, Masanori Ichihashi, Manabu Kubota, Chie Seki, Hitoshi Shimada, Yuhei Takado**: Collection of clinical data. **Kazunori Kawamura, Ming-Rong Zhang**: Radioligand synthesis. **Yoshiyuki Soeda, Akihiko Takashima**: Assay validation. **Makoto Higuchi, Takahiko Tokuda**: Conceptualization, Writing - review & editing, Funding acquisition, Supervision

