## Supplementary Methods and Results for "Development and Performance Assessment of a Novel Plasma p-Tau181 Assay Reflecting Tau Tangle Pathology in Alzheimer’s Disease"

#### 1. A new assay for phosphorylated tau protein (p-tau) in human plasma, which can detect both N- and C-terminally truncated p-tau181 (mid-p-tau181)

As shown in Supplementary Figure 1, an anti-tau mouse monoclonal antibody against a mid-portion of tau protein ranging from 151 to 163 amino acid residues was coupled to paramagnetic beads (Quanterix) and used for the capture antibody. As a detector, we used the AT270 mouse monoclonal antibody (Invitrogen) specific for the threonine 181 phosphorylation site. The detection antibody was conjugated to biotin following the manufacturer's instructions (Quanterix). We used the human p-tau181 standard in the Tau (Phospho) [pT181] Human ELISA Kit (Invitrogen) as the calibrator for our mid-p-tau181 assay. All plasma samples were diluted four times with the Tau Calibrator Diluent (Quanterix) prior to the assays to minimize matrix effects. The assay procedure followed that for the pTau-181 Advantage V2.1 Kit, except for changes to the capture antibody used as mentioned above. The details of the assay procedures have been described previously (Shinomoto et al. 2021; Satoh-Asahara et al. 2022). All plasma samples were run in duplicate with the same lot of standards. Fluorescent signals were converted to average enzyme per bead (AEB) numbers as described previously (Tatebe et al. 2017) and then concentrations of plasma mid-p-tau181 were extrapolated from four-parametric logistic curves of AEBs generated with known calibrator concentrations.

Plasma mid-p-tau181 levels were measured using the previously described assay on the Simoa HD-X Analyzer (Quanterix) at the National Institutes for Quantum Science and Technology. To ensure that measurements were performed in a blinded manner, unique identifiers that did not contain any participant-related information were attached to the blood sample tubes sent from the clinical department to the investigator who measured the plasma mid-p-tau levels. The samples were decoded only after data sharing with the participating clinical scientists had been completed. Internal quality control samples were analyzed in duplicates at the start and end of each run to determine within- and between-run variations.

#### 2. Validation of the newly developed mid-p-tau181 assay

##### 2-1. Standard curve for the mid-p-tau181 immunoassay

Supplementary Figure 2 shows the standard curve for our novel mid-p-tau181 assay, demonstrating that mid-p-tau181 was detected with high sensitivity. The curve was generated by analyzing the p-tau181 standard in the Tau (Phospho) [pT181] Human ELISA Kit (Invitrogen) in duplicate measures and fitting the digital signals on a fourparameter logistic curve. The goodness of fit was 0.9999.

### 2-2. Limit of detection (LOD) and lower limit of quantification (LLOQ)

We prepared 16 aliquots of blank sample (Tau Calibrator Diluent) and measured "background" signals of our novel plasma mid-p-tau181 assay on the Simoa HD-X Analyzer (Quanterix). Signals measured on the Simoa Analyzer were quantified by a common unit, namely average number of enzymes labels per bead (AEB). Thereafter, the LOD of the assay was determined as an interpolated mid-p-tau181 concentration derived from the mean plus 2.5 SD value of AEBs for the blank samples. The LOD of the assay, which requires 50 μL of plasma, was 0.1144 pg/mL.

The LLOQ of the assay was determined as an interpolated p-tau concentration derived from the mean plus 10 SD value of AEBs for blank samples. The LLOQ of the assay was 0.3770 pg/mL.

### 2-3. Intra-assay precision

Twenty samples with different concentrations (0.039, 6.25, 25, and 100 pg/mL) of recombinant p-tau181 were prepared for analysis of intra-assay precision, and measured AEBs in one experiment. Intra-assay precision was determined by calculating within-run coefficient of variation (CV) for those samples.

Intra-assay precision was robust with CVs between 1.1% and 3.5 % (Supplementary Table 1).

### 2-4. Inter-assay precision for quality controls and repeatability of the standard curve

We prepared three recombinant p-tau samples and two plasma samples with different plasma mid-p-tau181 concentrations for quality control experiments and measured the mid-p-tau181 levels in those samples five times on different days. Inter-assay precision was determined by calculating the CV of the AEB signals between the runs for those samples.

The %CV of the AEBs at different concentrations were 5.56, 5.09, 9.19, 5.30, and 5.96 %, respectively (Supplementary Table 2). These results indicated that our novel mid-p-tau181 assay showed good inter-assay precision (< 10%).

### 2-5. Dilution linearity

To explore dilution linearity, a plasma sample with a moderate mid-p-tau181 concentration was used, and twofold serial dilutions (×2, ×4, ×8, ×16, and ×32) of the sample were generated using the sample diluent until the theoretical concentration reached the LLOQ. Serially diluted samples were analyzed in duplicate.

The dilution linearity experiments demonstrated that an examined sample can be diluted to a concentration just below the LLOQ and still provide a reliable quantification after the serial dilution (Supplementary Figure 3).

**2-6. Spike recovery and parallelism**

For the spike recovery tests, four aliquots of each two plasma samples with different mid-p-tau181 concentrations were prepared and spiked with 0, 0.15, 0.3, and 0.6 pg of mid-p-tau181 in 360 µL of solution (containing patient plasma, sample buffer, and spiked peptide). These eight (4 × 2) aliquots were analyzed in quadruplicate on the same run. To evaluate parallelism, two spike recovery curves were made, starting from the non-spiked solution to the 0.6 pg-spiked solution. Recovery rates (%Recovery) were calculated by subtracting the endogenous mid-pTau181 concentration from the measured concentration (Andreasson U, et al. Front Neurol. 2015; 6: 179).

In the spike recovery and parallelism experiments (Supplementary Figure 4), the recovery rate (%Recovery) of each sample was 84.4%–119.3% (Supplementary Table 3). The parallelism of two spike recovery curves starting from the non-spiked solution to the 0.6 pg-spiked solution is also shown in Supplementary Figure 4, with our data demonstrating that plasma samples spiked with 0, 0.15, 0.3, and 0.6 pg of mid-p-tau181 protein provided reliable recovery rates and parallelism.

**2-7. Comparison of AEB signals for N- and C-terminally truncated p-tau181 between the established p-tau181 assay and the new mid-p-tau181 assay developed herein**

To confirm the ability of our new mid-p-tau assay to detect both N- and C-terminally truncated p-tau proteins, we compared our assay with the widely used and commercially available p-tau181 assay (Simoa™ pTau-181 Advantage V2 Kit, Quanterix). We prepared both N- and C-terminally truncated p-tau proteins as described in the following sections.

**2-7-1. cDNA construction**

The cDNA of tau (aa100−251) was generated from NΔR cDNA in PCI-neo (Takashima et al. 1998) according to the modified instructions of the PrimeSTAR® Mutagenesis Basal Kit (Takara Bio Inc., R046). Briefly, 2.5 ng of NΔR cDNA was mixed in 1× PrimeSTAR® Max DNA Polymerase (Takara Bio Inc., R045) containing 0.2 µM of forward primer (5′-CACCATGGGAACCACAGCTGAAGAAGCAGGC-3′) and 0.2 μM of reverse primer (5′-GTGGTTCCCATGGTGGCGAATTCTCGAGGCTAG-3′). PCR was performed on a C1000™ Thermal Cycler (BIO-RAD) with the following steps: an initial denaturation at 98°C for 2 min; 18 three-step cycles involving denaturation at 98°C for 10 s, annealing at 55°C for 15 s, and extension at 68°C for 90 s; and a final extension at 68°C for 10 min. A PCR product was incubated with Dpn I (20 units) (New England Biolabs, R0176) at 37°C for 20 min to digest the template NΔR plasmid. cDNA of 100−251 tau was transformed into DH5α (TOYOBO, DNA-903F). After the DH5α was cultured in LB medium containing 10 mg/mL of tryptone (Gibco, 211705), 5 mg/mL of yeast extract (Gibco, 212750), 10 mg/mL of sodium chloride (Nacalai Tesque, 31320-05), and 100 μg/mL of ampicillin (Nacalai Tesque, 02739-32), the cDNA was purified by NucleoSpin® Plasmid Transfection-grade (Takara Bio Inc., 740490). The sequences of the cDNA were confirmed through DNA sequencing (FASMAC).

**2-7-2. Cell culture**

COS-7 cells at around 90% confluence were detached from 10-cm dishes (CORNING, 353003) by trypsinization (Nacalai Tesque, 32777-15) and counted. The cells were plated at $2.0 \times 10^5$ in a 6-well plate (CORNING, 3506). cDNAs (2.5 μg) were transfected with Lipofectamine 3000 according to the manufacturer's instruction (Invitrogen, L3000) 2 days after culturing. PCI-neo (Promega) was used as the empty vector. Transfection reagents were removed by a medium change 1 day after the transfection.

**2-7-3. Preparation of cell homogenate**

The cells were scraped in a buffer (1 × TBS [Nacalai Tesque, 35438-81], 1 mM EDTA [Nacalai Tesque, 15112-22]) containing protease inhibitors (5 μg/mL of pepstatin A [Nacalai Tesque, 26436-52, 5 μg/mL of leupeptin [Nacalai Tesque, 103476-89-7], 2 μg/mL of aprotinin [Nacalai Tesque, 03346-84], and phosphatase inhibitors [Nacalai Tesque, 07575-51]) and homogenized for 30 s on the crashed ice using a polytron homogenizer (NITI-ON, NS-310E). After homogenization, solutions were centrifuged (23000 rpm, 15 min, 4°C) using a TLA55 Rotor (Beckman Coulter). The supernatants were applied to the Bradford assay (Nacalai Tesque, 11617-71) to determine the total protein concentration and then stored at −80 °C.

**2-7-4. Comparison between our mid-p-tau181 assay and the commercially**

**available p-tau181 assay**

Serially diluted cell homogenates, prepared as previously described, containing both N- and C-terminally truncated p-tau181 were applied to our mid-p-tau181 assay (Supplementary Figure 5, a solid line) and a commercially available p-tau181 assay (Simoa™ pTau-181 Advantage V2 Kit, Quanterix). Fluorescent signals converted to AEB values obtained from both assays are plotted in the Supplementary Figure 5 (solid line: mid-p-tau assay; broken line: commercially available p-tau181 assay). The results from both assays demonstrated that AEB values were obtained in our mid-p-tau181 assay but not in the commercially available p-tau181 assay, indicating that our mid-p-tau assay can quantify both N- and C-terminally truncated p-tau181 that could not be detected with the conventional p-tau181 assay.

**3. Determining the cutoff values of temporal meta-ROI SUVR in tau PET images**

ROC analysis was conducted using tau PET images of CN subjects and AD continuum patients to determine the optimal cutoff value for the temporal meta-ROI SUVR. Our analysis provided an AUC of 0.924. Moreover, based on the Youden index, we obtained a sensitivity and specificity of 85.4% and 92.5%, respectively. The optimal cutoff value for discrimination was determined to be 1.105.


Satoh-Asahara N, Yamakage H, Tanaka M, Kawasaki T, Matsuura S, Tatebe H, Akiguchi I, Tokuda T. Soluble TREM2 and Alzheimer-related biomarker trajectories in the blood of patients with diabetes based on their cognitive status. Diabetes Res Clin Pract. 2022; 193: 110121.

Shinomoto M, Kasai T, Tatebe H, Kitani-Morii F, Ohmichi T, Fujino Y, Allsop D, Mizuno T, Tokuda T. Cerebral spinal fluid biomarker profiles in CNS infection associated with HSV and VZV mimic patterns in Alzheimer's disease. Transl Neurodegener. 2021; 10(1): 2.

Tatebe H, Kasai T, Ohmichi T, Kishi Y, Kakeya T, Waragai M, Kondo M, Allsop D, Tokuda T. Quantification of plasma phosphorylated tau to use as a biomarker for brain Alzheimer pathology: pilot case-control studies including patients with Alzheimer's disease and down syndrome. Mol Neurodegener. 2017; 12(1): 63.

Takashima A, Murayama M, Murayama O, Kohno T, Honda T, Yasutake K, Nihonmatsu N, Mercken M, Yamaguchi H, Sugihara S, Wolozin B. Presenilin 1 associates with glycogen synthase kinase-3beta and its substrate tau. Proc Natl Acad Sci U S A. 1998; 95(16): 9637-41.

**Supplementary Table 1. Intra-assay precision (n = 20).**

| | Sample 1: 0.039 pg/mL | Sample 2: 6.25 pg/mL | Sample 3: 25.0 pg/mL | Sample 4: 100 pg/mL |
|---|---|---|---|---|
| Mean AEB | 0.00632 | 0.0580 | 0.224 | 0.872 |
| SD | 0.000222 | 0.000849 | 0.00656 | 0.00940 |
| CV (%) | 3.51 | 1.46 | 2.94 | 1.08 |

AEB, average enzyme per bead; SD, standard deviation; CV, coefficient of variation.

**Supplementary Table 2. Inter-assay precision for quality control samples (n = 25).**

| | Sample 1 | Sample 2 | Sample 3 | Sample 4 | Sample 5 |
|---|---|---|---|---|---|
| Mean AEB | 0.00331 | 0.00484 | 0.0592 | 0.128 | 0.882 |
| SD | 0.000184 | 0.000246 | 0.00545 | 0.00676 | 0.0526 |
| CV (%) | 5.56 | 5.09 | 9.19 | 5.30 | 5.96 |

AEB, average enzyme per bead; SD, standard deviation; CV, coefficient of variation.

**Supplementary Table 3. Recovery rate (%Recovery) for each plasma sample in the spike recovery tests.**

| | Treatment | Measured concentration (pg/mL) | CV (%) | Theoretical concentration of the spiked sample (pg/mL) | Recovery rate (%) |
|---|---|---|---|---|---|
| Sample 1 | Neat (0 pg spike) | 0.2675 | 14.9 | | |
| | 0.15 pg spike | 0.3500 | 7.94 | 0.4175 | 119.3 |
| | 0.3 pg spike | 0.5130 | 12.2 | 0.5675 | 110.6 |
| | 0.6 pg spike | 0.8649 | 7.73 | 0.8675 | 100.3 |
| Sample 2 | Neat (0 pg spike) | 0.4516 | 6.91 | | |
| | 0.15 pg spike | 0.6842 | 3.63 | 0.6016 | 87.9 |
| | 0.3 pg spike | 0.8909 | 5.82 | 0.7516 | 84.4 |
| | 0.6 pg spike | 1.101 | 7.44 | 1.0516 | 95.6 |

CV, coefficient of variation.

**Supplementary Table 4. Multiple linear regression analysis of p-tau levels with amyloid and tau PET metrics.**

| | Beta coefficient | Standard error | Adjusted $R^2$ | P-value |
|---|---|---|---|---|
| mid-p-tau181 | | | | |
| Amyloid PET (Centiloid) | 0.399 | 0.118 | 0.420 | 0.002 |
| Tau PET (Temporal meta-ROI) | 0.311 | 0.118 | | 0.020 |
| N-p-tau181 | | | | |
| Amyloid PET (Centiloid) | 0.258 | 0.115 | 0.450 | 0.056 |
| Tau PET (Temporal meta-ROI) | 0.472 | 0.115 | | <0.0001 |

**Supplementary Table 5. Tau PET status of cognitively normal individuals and AD continuum patients through three different approaches.**

| | CN | | AD | |
|---|---|---|---|---|
| Quantitative approach | Tau (-) | Tau (+) | Tau (-) | Tau (+) |
| Braak staging | 35 | 5 | 3 | 45 |
| Temporal meta-ROI SUVR | 37 | 3 | 7 | 41 |
| AD tau score | 40 | 0 | 2 | 35 |

The cut-off values of Temporal meta-ROI SUVR and AD tau score were 1.105 and 0.1986, respectively. AD tau scores were exclusively computed for cases that underwent imaging with the mCT PET scanner.

**Legends for Supplementary Figures**

**Supplementary Figure 1. A schematic illustration tau protein showing the epitope location of the antibodies used in this study.**

**Supplementary Figure 2. Standard curve.**
Refer “**Supplementary Methods and Results**”, “**2-1. Standard curve for the mid-p-tau181 immunoassay**”.

**Supplementary Figure 3. Dilution linearity.**
Refer “**Supplementary Methods and Results**”, “**2-5. Dilution linearity**”.

**Supplementary Figure 4. Spike recovery and parallelism.**
Refer “**Supplementary Methods and Results**”, “**2-6. Spike recovery and parallelism**”.

**Supplementary Figure 5. Comparison of the AEB signals between our mid-p-tau181 assay and the commercially available p-tau181 assay.**
Refer “**Supplementary Methods and Results**”, “**2-7-4. Comparison between our mid-p-tau181 assay and the commercially available p-tau181 assay**”.

**Supplementary Figure 6. List of ROIs applied to the tau PET images.**
From left to right, overlaid ROIs on native T1 space of representative subject: Braak staging ROI, temporal meta-ROI, and AD tau score ROI. The Braak staging ROI is color-coded, with Braak I/II, III/IV, and V/VI regions shown in red, yellow, and blue, respectively. The temporal meta-ROI is labeled in green. The AD tau score ROI represents 114 areas segmented using Mvision. Scores were derived by assigning weights to diagnostically relevant areas within these ROIs.

**Supplementary Figure 7. Comparative analyses of blood biomarkers across the clinical diagnosis groups and between the amyloid PET status.**
(A) The data distribution is portrayed using scatter plots, with group differences having been evaluated through the Kruskal–Wallis test. Statistically significant differences are denoted as follows: $p < 0.0001$ (****), $p = 0.0004$, or 0.0010 (***), $p = 0.0070$ (**), and non-significant at $p < 0.05$ (ns), as determined using Dunn's multiple comparisons.
(B) ROC curves were utilized to differentiate AD from other groups determined according to qualitative findings of amyloid PET. The corresponding AUC values are

listed.
CN, cognitively normal; AD, Alzheimer’s disease; PSP, progressive supranuclear palsy; FTLD, frontotemporal degeneration; Aβ, amyloid beta; N-p-tau181, phosphorylated tau181 measured with the Simoa pTau-181 V2.1 Assay kit (Quanterix); mid-p-tau181, phosphorylated tau181 measured using the originally developed immunoassay directed to both N- and C-terminally truncated p-tau181 fragments; NfL, neurofilament light chain; ROC, Receiver Operating Characteristic; AUC, area under the ROC curve.

**Supplementary Figure 8. Correlations between plasma mid-p-tau levels and tau PET measures in the subjects with PSP.**
Correlation analysis between tau PET images and plasma mid-p-tau181 levels in patients with PSP. Neither voxel-based (A) nor ROI-based (B and C) analyses showed a significant correlation with tau PET accumulation. The ROI-based approach used Pearson's correlation, with the results presented as r (correlation coefficient) and p (statistical significance) values. PSP-tau score is a machine learning-based measure indicating PSP-related features of tau PET images (Endo et al. 2022). Each anatomical ROI was set in MNI space using a template atlas (Talairach Daemon atlas from the Wake Forest University PickAtlas version 3.0.5).
SUVR, standardized uptake value ratio.
