## Supplementary Figures for "Development and Performance Assessment of a Novel Plasma p-Tau181 Assay Reflecting Tau Tangle Pathology in Alzheimer’s Disease"

### Supplementary Figure 1

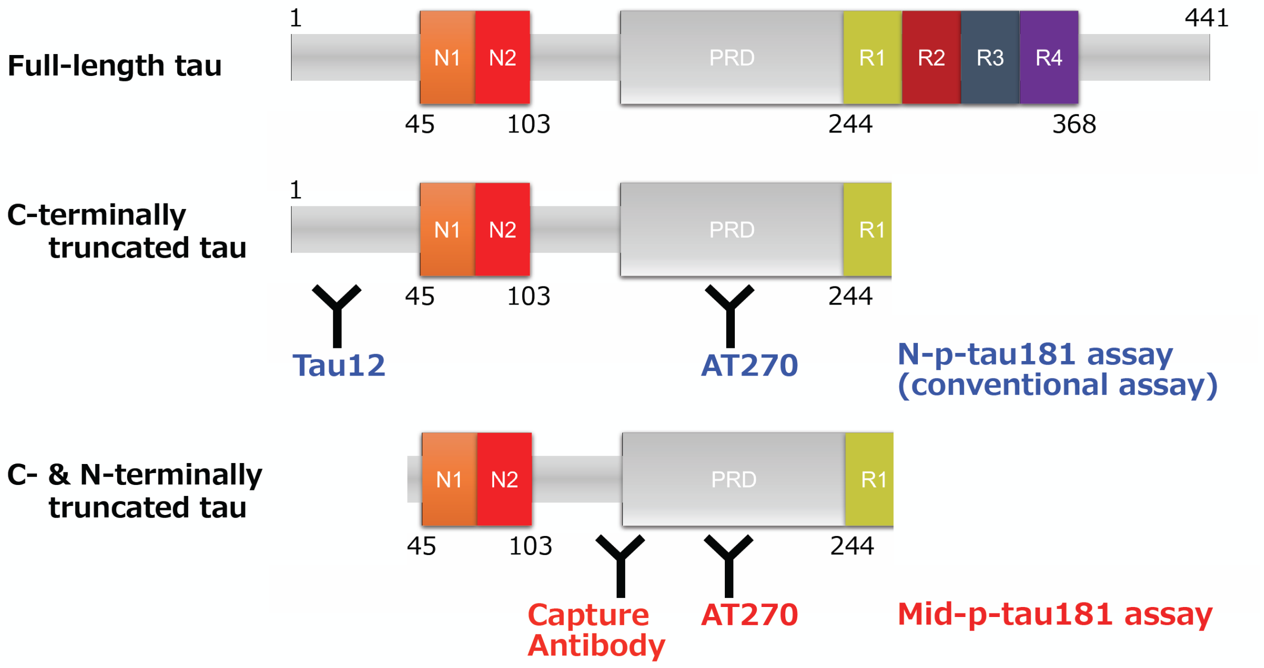

### Supplementary Figure 2

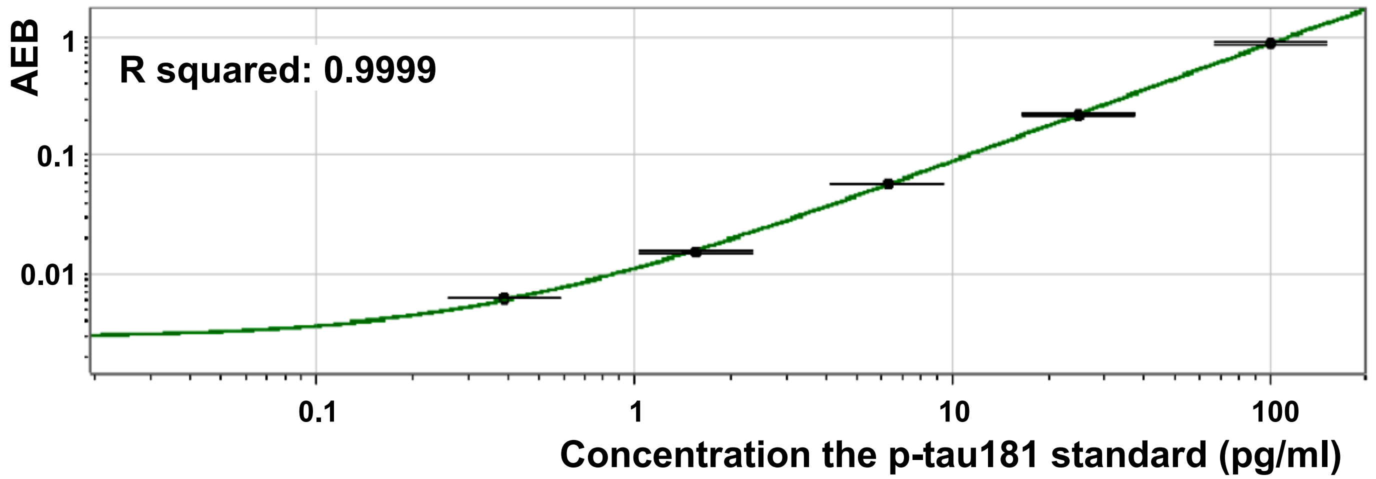

### Supplementary Figure 3

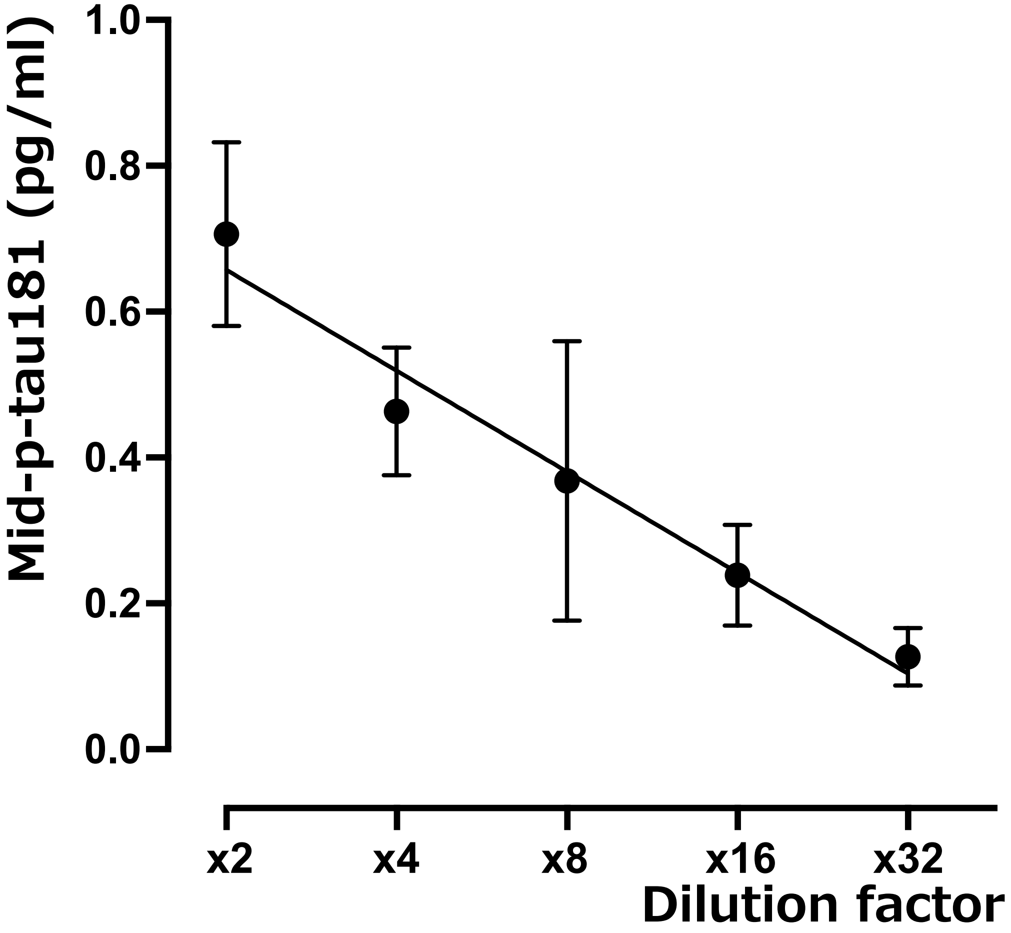

### Supplementary Figure 4

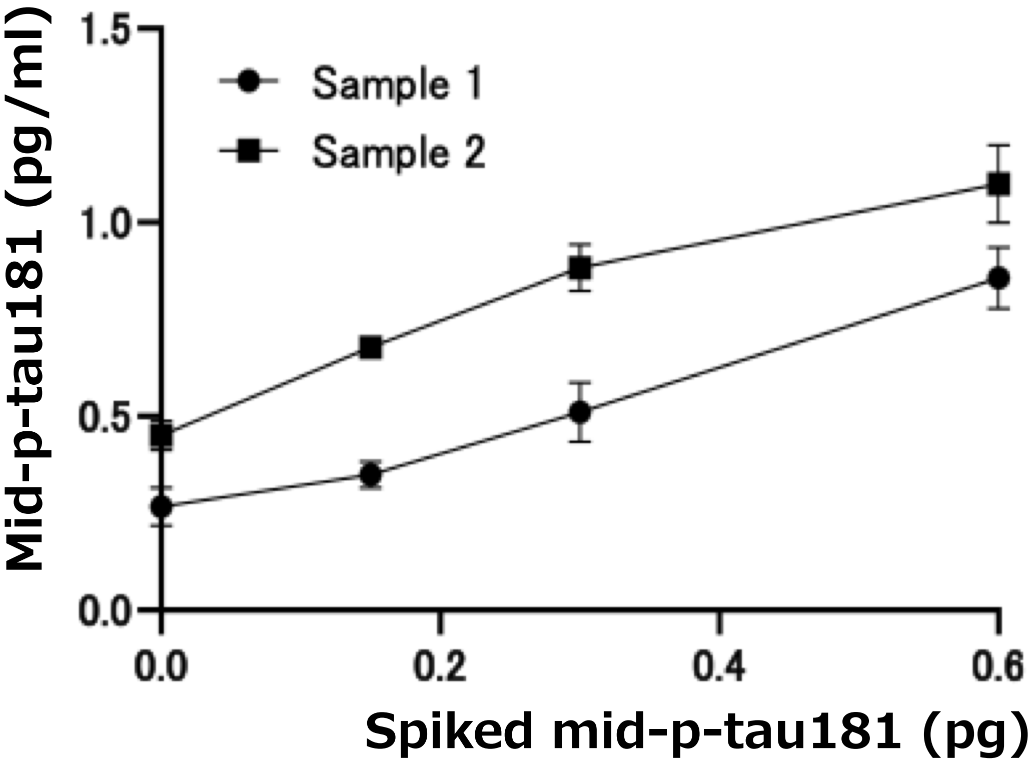

### Supplementary Figure 5

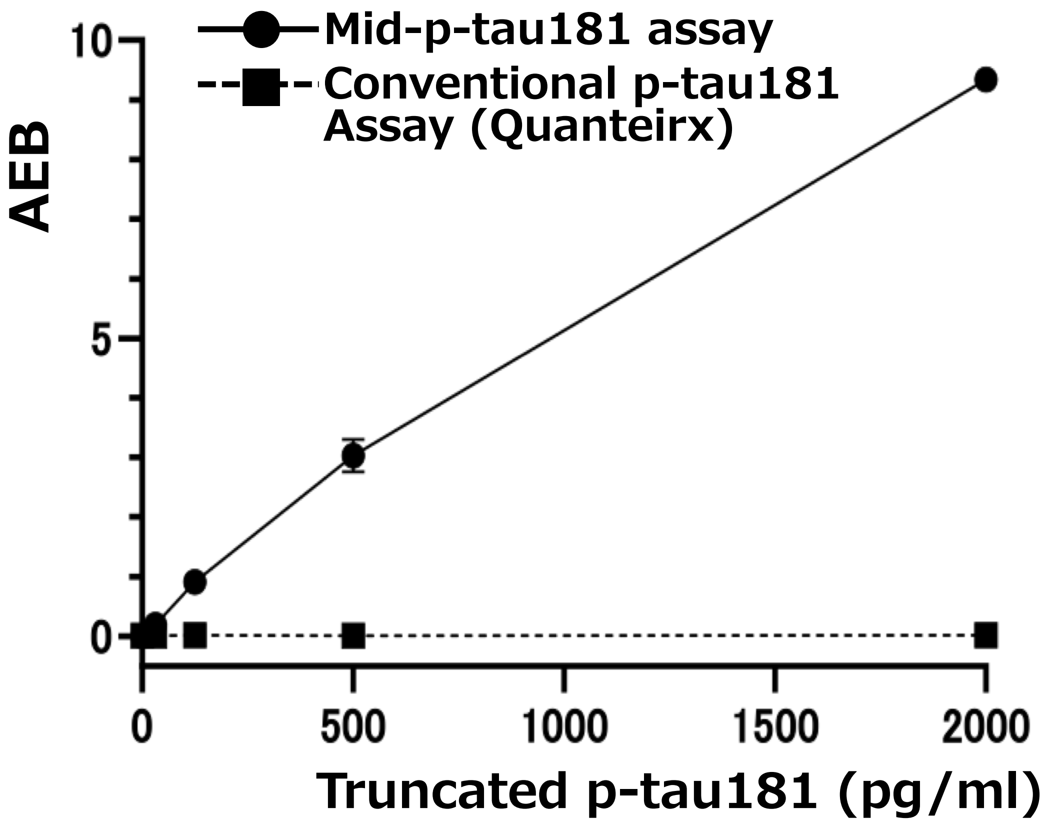

### Supplementary Figure 6

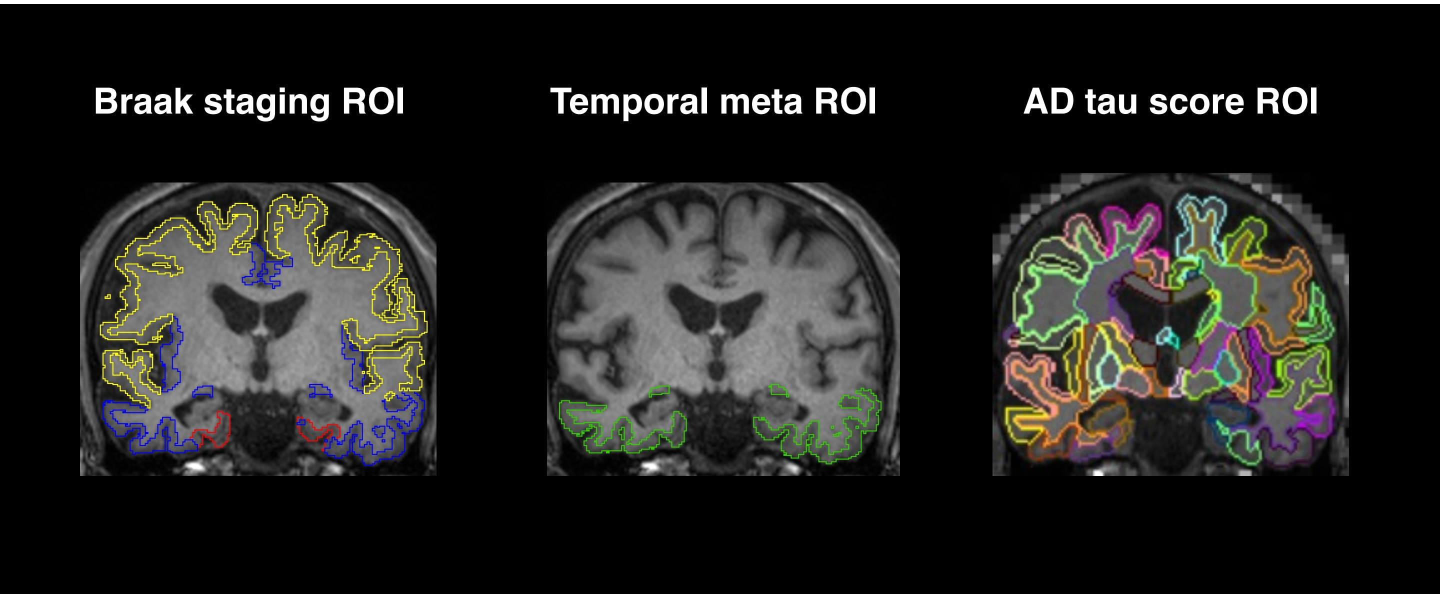

### Supplementary Figure 7

(A)

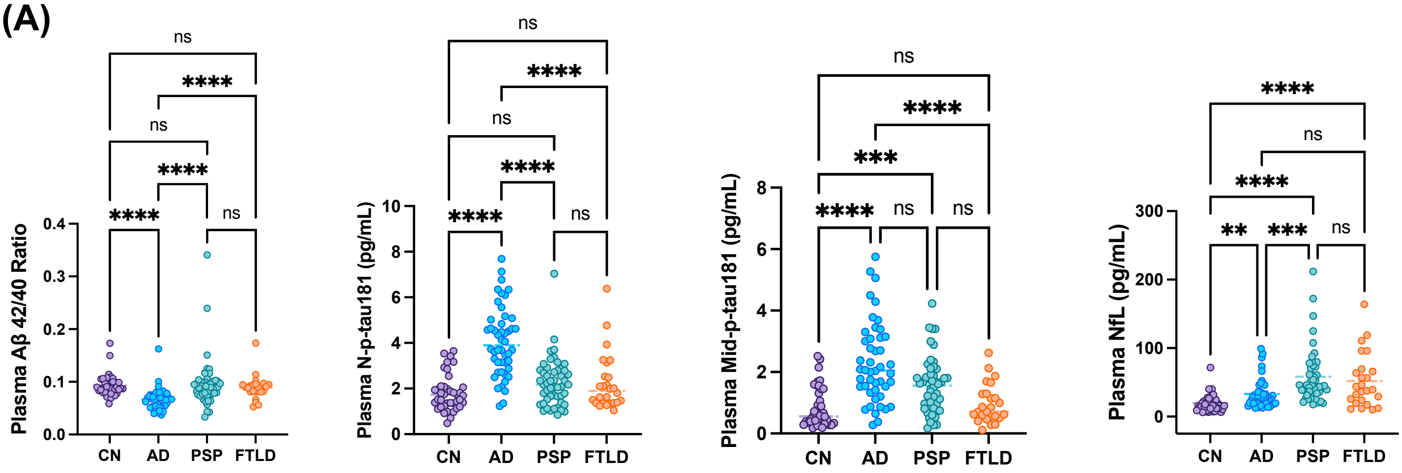


(B) Amyloid (+) vs Amyloid (-)

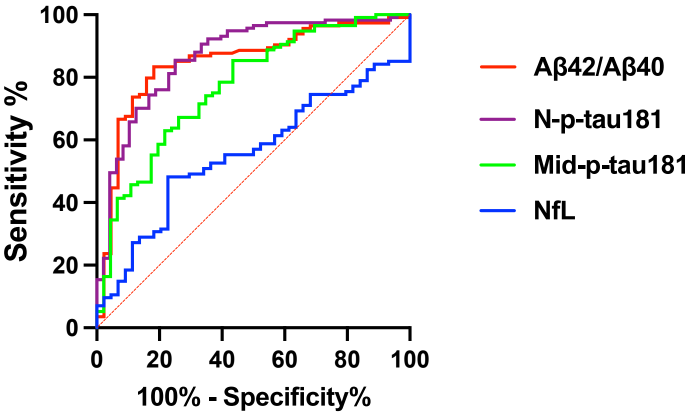


| | AUC values |
|---|---|
| Aβ42/40 | 0.714 |
| N-p-tau181 | 0.905 |
| Mid-p-tau181 | 0.844 |
| NfL | 0.641 |

### Supplementary Figure 8

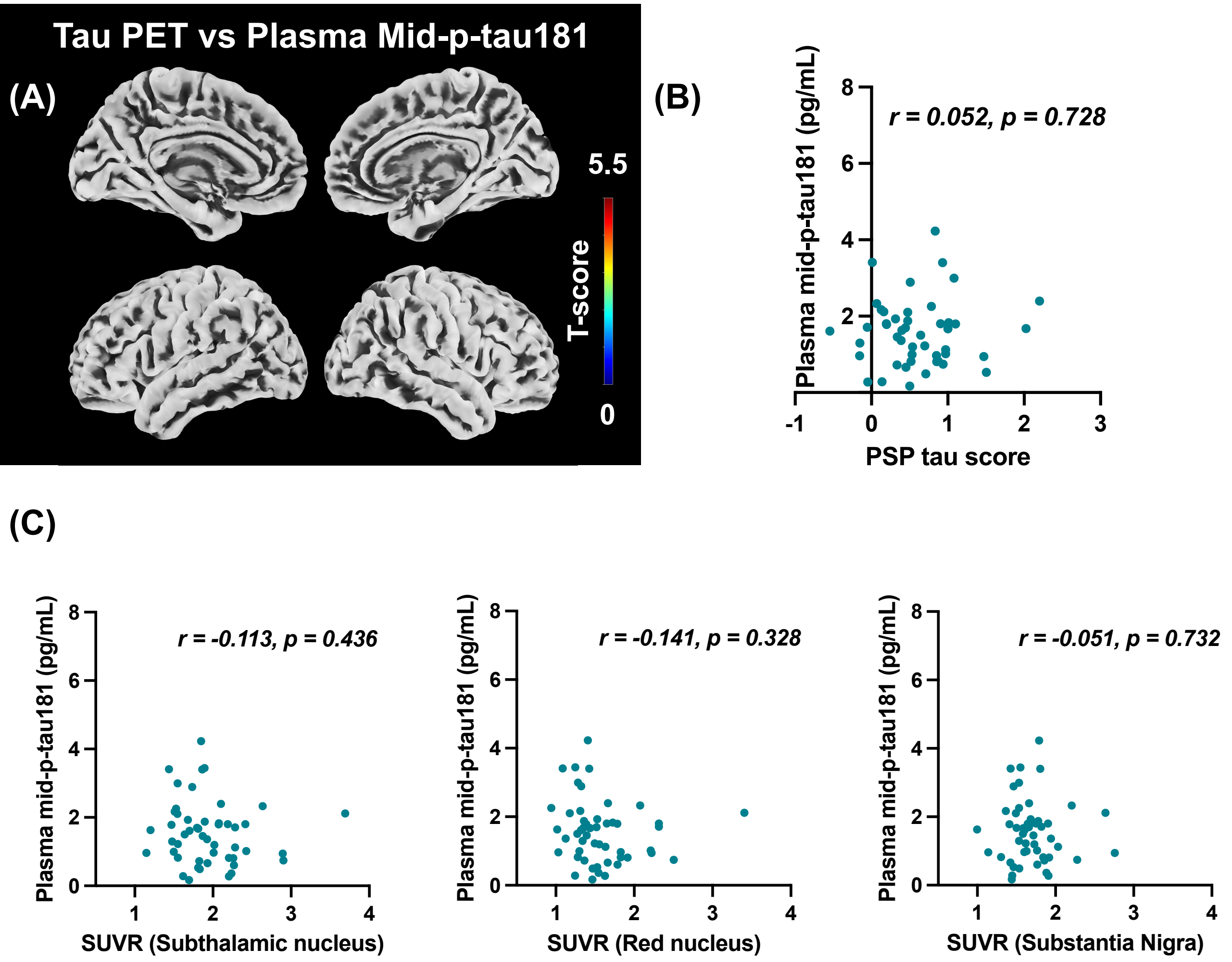
